# Pathogenic accumulation of T follicular helper cells in lupus disease depends on PD-L1 and IL-4 expressing basophils

**DOI:** 10.1101/2023.01.10.23284399

**Authors:** John Tchen, Quentin Simon, Léa Chapart, Yasmine Lamri, Fanny Saidoune, Emeline Pacreau, Christophe Pellefigues, Julie Bex-Coudrat, Hajime Karasuyama, Kensuke Miyake, Juan Hidalgo, Padraic G. Fallon, Thomas Papo, Ulrich Blank, Marc Benhamou, Guillaume Hanouna, Karim Sacre, Eric Daugas, Nicolas Charles

## Abstract

Systemic lupus erythematosus (SLE) is an autoimmune disease characterized by autoantibodies raised against nuclear antigens and whose production is promoted by autoreactive T follicular helper (TFH) cells. Basophils, by accumulating in secondary lymphoid organs (SLO), amplify autoantibody production and disease progression through mechanisms to be defined. Here, we demonstrate that a functional relationship between TFH cells and basophils occurs in SLO during lupus pathogenesis. On SLE patient blood basophils, PD-L1 expression was upregulated and associated with TFH and TFH2 cell expansions and with disease activity. In two distinct lupus-like mouse models, TFH cell pathogenic accumulation, maintenance and function, and disease activity were dependent on basophils and their expressions of PD-L1 and IL-4. Our study establishes a direct link between basophils and TFH cells in the SLE context that promotes autoreactive IgG production and lupus nephritis pathogenesis. Altering the basophil/TFH cell axis in the SLE context may represent a promising innovative intervention strategy in SLE.

## INTRODUCTION

Systemic lupus erythematosus (SLE) is a multifactorial autoimmune disease that can affect different organs including joints, skin, or kidneys (lupus nephritis). A break in tolerance to nuclear self-antigens leads to an accumulation of autoreactive B and T cells in SLE patients that drives the production of autoreactive antibodies mainly raised against nuclear antigens, such as double-stranded DNA (dsDNA) or ribonucleoproteins (RNP). These autoantibodies form circulating immune complexes (CIC) after aggregation to complement factors and autoantigens. CIC deposit in target organs where they can induce a chronic inflammation leading to tissue damage and organ dysfunction. In parallel, CIC activate innate immune cells such as plasmacytoid dendritic cells, monocytes/macrophages, neutrophils, and basophils that enhance autoantibody production through the release of various inflammatory mediators and initiate a deleterious amplification loop of the disease^1,2^.

Autoantibody production is a key event in lupus pathogenesis. Its disruption represents an intensive area of clinical development of biotherapies targeting directly the B cells or pathways promoting their survival and maturation^3^. Recent advances in the understanding of SLE pathogenesis have shed some light on the role of T follicular helper CD4^+^ (TFH) cells. TFH cells are central for follicular B cell maturation into antibody-secreting cells and their numbers and functions are dysregulated in both human SLE patients and some lupus-like murine models^4-8^. TFH cells are characterized by their expression of the transcription factor BCL6 and their surface expression of C-X-C motif chemokine receptor 5 (CXCR5) which allows their localization into the germinal centers to provide B cell help through IL-21 production and promote the maturation of antibody-secreting cells^7^. TFH cells express programmed cell-death 1 receptor (PD-1) that is essential for their positioning, functions, and regulation in secondary lymphoid organs (SLO)^9^. GATA-3 transcription factor expression, IL-4 production, and potent help provided to B cells characterize the TFH type 2 (TFH2) cell subset that is overrepresented in the lupus context and associated with disease activity^6^.

Basophils are activated during SLE pathogenesis and accumulate in SLO where they promote autoantibody production by supporting antibody-secreting plasmablast accumulation^10,11^. Depleting basophils in lupus-like mouse models with established disease dampens lupus-like activity by reducing plasmablast numbers, autoantibody titers, CIC deposits in glomeruli, and kidney inflammation^10,11^, thus demonstrating that basophils are responsible for an amplification loop driving the disease to a pathogenic threshold. In agreement, constitutive basophil deficiency prevents lupus-like disease onset in the pristane-induced lupus-like disease model^12^. In this context, targeting the addressing of basophils to SLO has demonstrated promising therapeutic potential^11,13^. In the *Lyn*^*–/–*^ lupus-like mouse model, we previously showed that basophils accumulate in SLO where they express surface molecules that suggest interactions with the B and T cell compartments to promote plasma cell maturation and antibody secretion^10^. However, the mechanisms by which basophils, once in SLO, actually promote this disease amplification loop remains unknown.

Here, we screened cell surface molecules on blood basophils from SLE patients that could support a functional interaction of these cells with SLO cell partners in the lupus context. We demonstrate *in vitro* in humans and *in vivo* in two different SLE mouse models that PD-L1 expression and IL-4 production by basophils are required to promote the cross-talk of these cells with TFH cells to sustain TFH cell pathogenic accumulation, TFH2 cell differentiation, and thereby mediate SLE disease onset and progression.

## RESULTS

### Human blood basophils from SLE patients overexpress PD-L1

TFH, and especially TFH2, cells are significant contributors to the physiopathology of SLE. Proportions among CD4^+^ T cells of circulating TFH (cTFH) and cTFH2 cells are increased in several SLE patient cohorts and lupus-like mouse models^4-7,14,15^. We first assessed these cTFH cells (CD3^+^ CD4^+^ CXCR5^+^ ICOS^+^ PD-1^+^ cells) and cTFH2 cells (CCR6^−^ CXCR3^−^ TFH cells) in our SLE patient cohort (**Table S1**) and confirmed both their increased representation among CD4^+^ T cells and a cTFH2 cell bias that was associated with disease activity (**Fig. 1a-c**).

**Figure 1:**
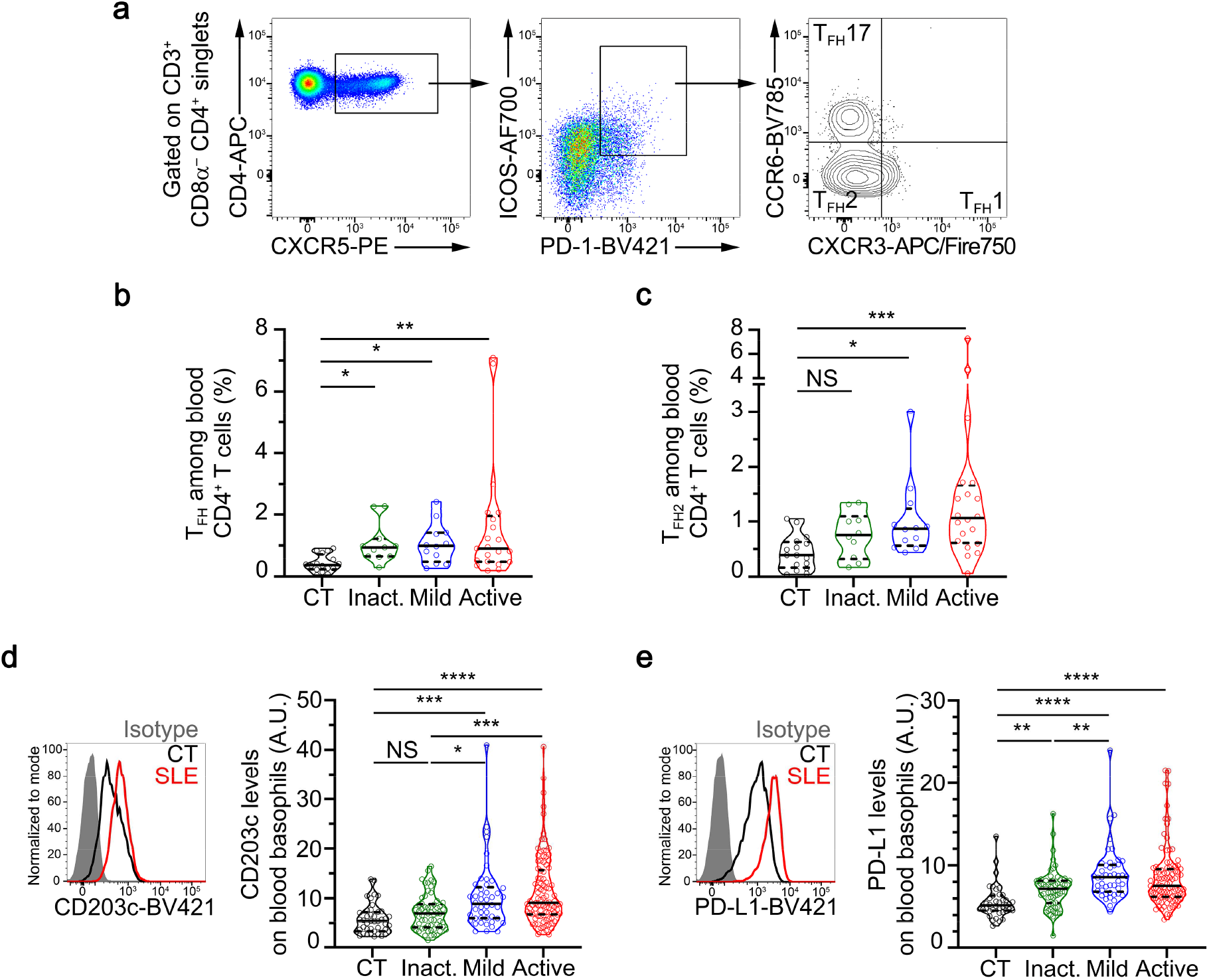
Human blood basophils from SLE patients overexpress PD-L1. (**a**) Flow cytometry gating strategy used to identify human circulating T follicular helper T cells (TFH) defined as CD3^+^ CD8α^−^ CD4^+^ CXCR5^+^ ICOS^+^ PD-1^+^. Among TFH cells, TFH1 cells were defined as CXCR3^+^CCR6^−^, TFH2 as CXCR3^−^ CCR6^−^, and TFH17 as CXCR3^−^ CCR6^+^. (**b**) Proportions (%) among CD4^+^ T cells of TFH as defined in (**a**) in blood from healthy controls (CT) and inactive (inact.), mild, or active SLE patients (n = 16/11/12/21, respectively) as determined by flow cytometry. (**c**) Proportions (%) among CD4^+^ T cells of TFH2 as defined in (**a**) in blood from healthy controls and inactive, mild, or active SLE patients (n = 16/10/12/20, respectively) as determined by flow cytometry. (**d**) **Left**, Histogram plot representing the CD203c expression levels on blood basophils from a healthy control (CT, black line), a patient with active SLE (red line), and isotype control staining (grey filled histogram). **Right**, CD203c expression levels on blood basophils from CT and inact., mild or active SLE patients (n = 43/61/46/98, respectively) as determined by flow cytometry. (**e**) **Left**, Representative histogram plot of PD-L1 expression levels on blood basophils as in (**d**). **Right**, PD-L1 expression levels on blood basophils from CT and inact., mild or active SLE patients (n = 39/60/45/96, respectively) as determined by flow cytometry. (**b**-**e**) Data are presented as violin plots with median (plain line) and quartiles (dotted lines). Statistical analyses were Kruskal-Wallis tests followed by Dunn’s multiple comparisons tests. NS: not significant, *P < 0.05, **P < 0.01, ***P < 0.001, ****P < 0.0001. A.U.: arbitrary units.

We and others reported that peripheral basopenia and activation of blood basophils correlate with disease activity in SLE patients^10,11,16^. Our current SLE patient cohort further confirmed this blood basophil phenotype as evidenced by basopenia and overexpression of the activation marker CD203c, the chemokine receptor CXCR4, and the L-selectin CD62L, all of them being associated with disease activity (**Fig. S1a-e** and **Fig. 1d**).

To investigate whether, in the SLE context, human basophils were more prone to interact with TFH cells, we assessed the expression levels of several surface markers identified to be relevant interacting molecules with TFH cells. PD-L2, ICOSL, and OX40L were barely detected on the surface of human blood basophils and no difference in their expression levels on basophils from healthy donors or SLE patients was observed (**Table S2**). Nevertheless, PD-L1 was strongly upregulated on the surface of SLE patient blood basophils as compared to healthy donor blood basophils, independently of the activity of the disease (**Fig. 1e** and **Table S2**). In addition, PD-L1 expression on SLE patient blood basophils correlated with the basophil activation status (Spearman r=0.3113, P < 0.0001, n=204) (**Fig. S1f**). CD84, a member of the SLAM family of proteins important for TFH cell regulation^17^, was highly expressed by human basophils (**Fig. S1g** and **Table S2**). Although no significant increase in CD84 expression on blood basophils from the whole SLE patient cohort was found, its expression was significantly increased on blood basophils from active SLE patients as compared to healthy volunteers (**Fig. S1g** and **Table S2**).

Altogether, these results demonstrated that basophils from SLE patients overexpress PD-L1 and CD84, suggesting that a basophil-TFH cell interaction may occur during lupus pathogenesis.

### Basophil-TFH cell axis in lupus-like mouse models

We next sought to verify whether this increased expression of PD-L1 on basophils from SLE patients was also detected on basophils from lupus-like mouse models in which basophil contribution to disease activity is established^10,11,18^. PD-L1 expression was increased on the surface of basophils from both pristane-induced and *Lyn*^*–/–*^ lupus-like mouse models in the analyzed compartments (blood, spleen, and lymph nodes) (**Fig. 2a,b**). As shown in other human studies and previous reports^7,11^, increased TFH cell proportions among CD4+ T cells and basophil accumulation were observed in both lupus-like mouse models in blood and the secondary lymphoid organs spleen, and lymph nodes (**Fig. 2c-f** and **Fig. S2**). These results suggest a positive correlation between basophils and TFH cells during lupus-like disease.

**Figure 2:**
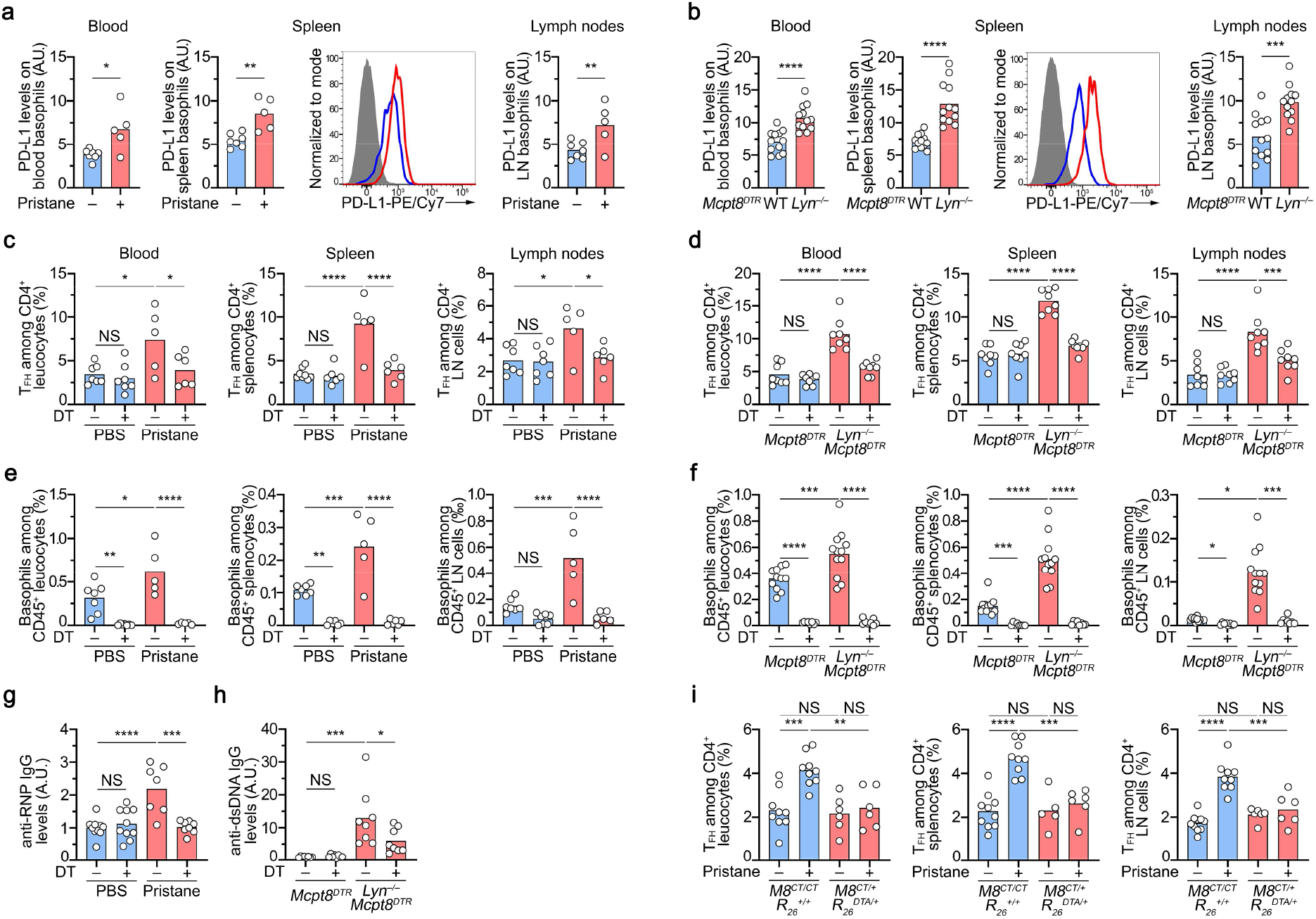
Basophil-TFH functional relationship during lupus-like disease. (**a**) PD-L1 expression levels on basophils from PBS-injected (blue bars) or pristane-injected *Mcpt8*^*DTR*^ mice (red bars) in the indicated compartments as determined by flow cytometry using the gating strategy described in **Fig. S2a**,**b**. A representative FACS histogram of PD-L1 expression on spleen basophils from a PBS-injected mouse (blue line), a pristane-injected mouse (red line), and the signal from the isotype control (grey filled) is shown. (**b**). PD-L1 expression levels on basophils from an aged *Mcpt8*^*DTR*^ (blue bars) or aged *Lyn*^*–/–*^ *Mcpt8*^*DTR*^ mice (red bars) in the indicated compartments as in (**a**). A representative FACS histogram of PD-L1 expression on spleen basophils from an aged *Mcpt8*^*DTR*^ (blue line), an aged *Lyn*^*–/–*^ *Mcpt8*^*DTR*^ mouse (red line), and the signal from the isotype control (grey filled) is shown. (**c**) Proportions (%) of TFH among CD4^+^ T cells in the indicated compartments from PBS-injected (blue bars) and basophil sufficient (DT–) or basophil-depleted (DT+) mice and from pristane-injected *Mcpt8*^*DTR*^ mice (red bars) DT treated or not. Data were determined by flow cytometry using the gating strategy described in **Fig. S2c**,**d**. (**d**) Proportions (%) of TFH among CD4^+^ T cells in the indicated compartments from aged *Mcpt8*^*DTR*^ (blue bars) and basophil sufficient (DT–) or basophil-depleted (DT+) mice and from aged *Lyn*^*–/–*^ *Mcpt8*^*DTR*^ mice (red bars) DT-treated or not. Data were determined by flow cytometry as in (**c**). (**e**) Proportions (%) of basophils in the indicated compartments in the same mice as described in (**c**) were determined by flow cytometry as in (**a**). (**f**) Proportions (%) of basophils in the indicated compartments in the same mice as described in (**d**) were determined by flow cytometry as in (**a**). (**g**) Anti-RNP IgG autoantibody plasma titers from the same mice as in (**c**) were quantified by ELISA, as described in the methods section. O.D. values at 450 nm were determined and data were normalized to the mean of PBS-injected DT – *Mcpt8*^*DTR*^ values. (**h**) Anti-dsDNA IgG autoantibody plasma titers from the same mice as in (**d**) were quantified by ELISA, as described in the methods section. O.D. values at 450 nm were determined and data were normalized to the mean of DT – *Mcpt8*^*DTR*^ values. A.U.: arbitrary units. (**i**) Proportions (%) of TFH among CD4^+^ T cells in the indicated compartments from basophil-sufficient (*Mcpt8*^*CT/CT*^ *R26*^*+/+*^) (blue bars) or basophil-deficient (*Mcpt8*^*CT/+*^ *R26*^*DTA/+*^) (red bars) mice treated with PBS or pristane (pristine – or +, respectively). Data were determined by flow cytometry as in (**c**). (**a-i**) Results are from at least three independent experiments and presented as individual values in bars representing the mean values. Statistical analyses were done by one-way ANOVA followed by Holm-Šídák’s multiple comparisons test between the indicated groups. NS: not significant, p>0.05; *: p<0.05; **: p<0.01; ***: p<0.001; ****: p<0.0001.

To evaluate whether a functional relationship between basophils and TFH cells was taking place during the course of the disease, we depleted basophils through diphtheria toxin injection in *Mcpt8*^*DTR*^ mice in both lupus-like models during ten days before their analysis (**Fig. S2a**,**b**). We previously showed that basophil depletion dampens CD19^+^CD138^+^ short-lived plasma cell numbers in SLO, autoantibody titers, kidney deposits and inflammation, and disease activity in these models^10,11,18^. Here, in agreement, efficient DT-mediated basophil depletion (**Fig. 2e,f** and **Fig. S2a**,**b**) resulted in a dramatic decrease in autoreactive IgG antibody titers in both pristane-induced and *Lyn*^*– /–*^ lupus-like mouse models (**Fig. 2g,h**). In addition, it abrogated the increase in TFH cell proportions in blood and SLO from lupus-like mice (**Fig. 2c,d** and **Fig. S2c,d**), strongly suggesting a functional relationship between the two cell compartments. Of note, basophil depletion in *Lyn*^*–/–*^ *Mcpt8*^*DTR*^ mice did not modify the proportion of regulatory TFH (TFR) cells among the TFH cell population (**Fig. S3a,b**).

We recently showed that basophils play a nonredundant role in pristane-induced lupus-like disease and that basophil-deficient mice (*Mcpt8*^*CT/+*^ *Rosa26*^*DTA/+*^ mice) are resistant to pristane-induced lupus-like disease onset^12^. The functional relationship between basophils and TFH cells was further confirmed in this mouse model since no increase in the TFH cell proportion was observed in pristane-injected basophil-deficient mice as compared to control mice (*Mcpt8*^*CT/CT*^ *Rosa26*^*+/+*^) in any of the analyzed compartments (**Fig. 2i**).

The functional interaction between the two cell types seemed restricted to the lupus-like environment. Indeed, basophil depletion in WT animals in both models or constitutive basophil deficiency did not modify the basal proportions of TFH cells in the observed compartments (**Fig. 2c,d,i**). Moreover, basophils, which are not involved in the humoral response to the ovalbumin (OVA) protein after intraperitoneal immunization in alum^19^, did not influence the rise in TFH cell numbers in these conditions. Indeed, we could measure the induction of TFH cells in OVA-immunized mice which was associated with the production of anti-OVA IgG antibodies but not with the accumulation of basophils in the spleen nor in the draining (mesenteric) lymph node (**Fig. S3c-f**). DT-mediated basophil depletion starting the last 48 hours of the immunization procedure did not modify the TFH cell response nor the anti-OVA IgG titers in immunized mice, suggesting that basophils were not involved in the support of TFH cells in a protein-immunization setting, unlike what we observed in the lupus-like context (**Fig. S3c-f** and **Fig. 2**).

Altogether, these data strongly suggest that basophils, by accumulating in SLO, enable the expansion of the TFH cell population in two distinct lupus-like mouse models and that basophils and TFH cells may share a functional relationship in the lupus-like context.

### Basophils control TFH cell ability to produce IL-21 and IL-4 in a lupus-like context

We next assessed whether basophils could influence the cytokine production abilities of the expanded TFH cells during the disease. First, the proportions of spleen TFH cells producing IL-21 without any restimulation were significantly increased in both lupus-like models and further amplified after phorbol myristate acetate (PMA) and ionomycin restimulation as compared to spleen TFH cells from control mice (**Fig. 3a-c**). This suggested that in the lupus-like context, TFH cells are more prone to provide IL-21 to surrounding cells in these mouse models. Second, the same observations were done concerning IL-4 production by TFH cells showing a TFH2 bias in these two lupus-like models (**Fig. 3d-f**), as was also observed in the SLE patient cohort (**Fig. 1c**). Of note, no such bias was observed for IFNγ production by TFH cells in any of the mouse models (**Fig. 3g,h**). Third, basophil depletion dramatically dampened both constitutive and PMA-ionomycin-induced IL-21 and IL-4 productions by TFH cells only in the lupus-like context, without significantly influencing their IFNγ production ability (**Fig. 3a-h**). These results strongly suggest that, in the lupus-like context, basophils promoted TFH cell-derived IL-21 and IL-4 production ability and were responsible for a TFH2 cell bias of this T cell population.

**Figure 3:**
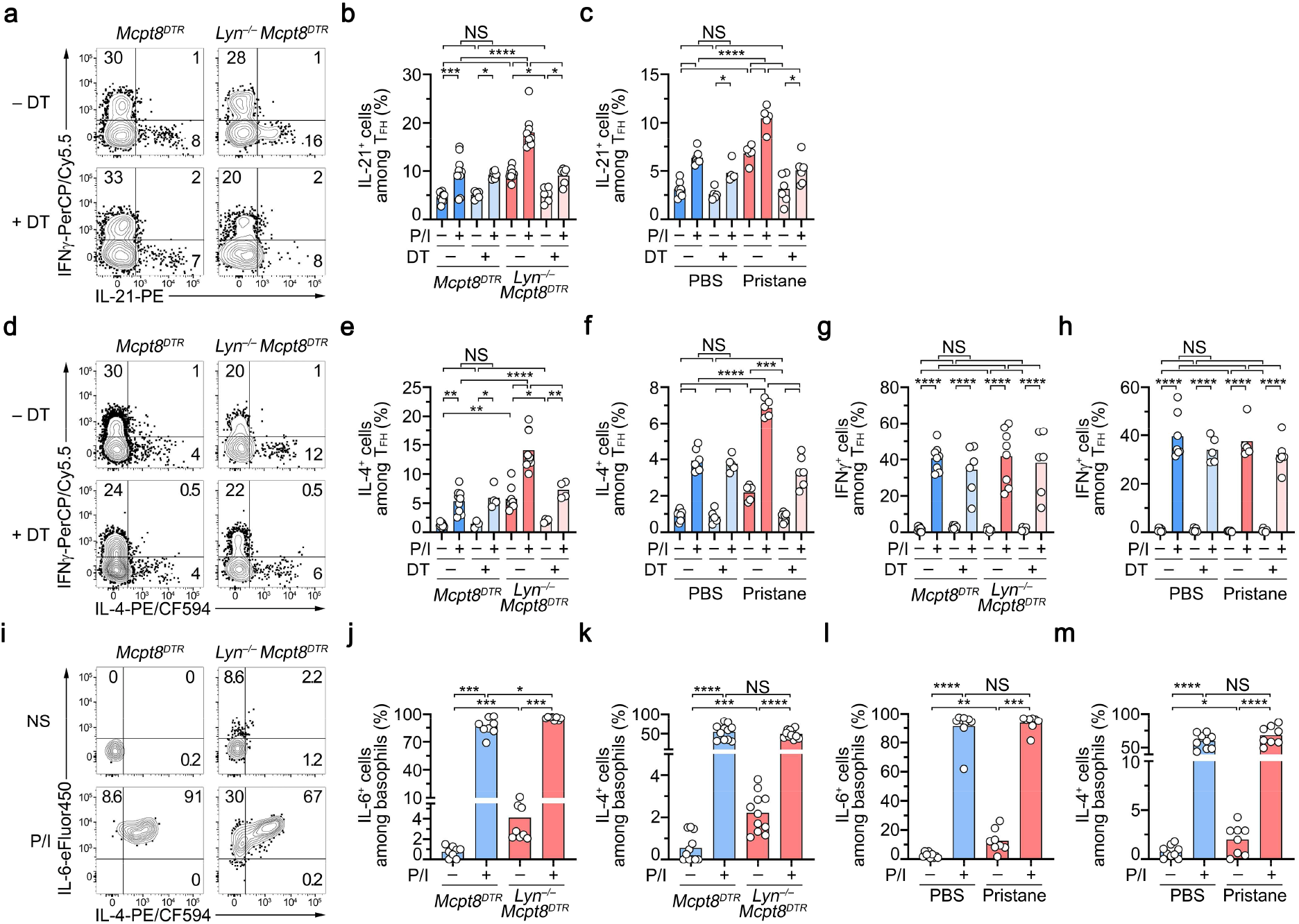
Basophils control TFH abilities to produce IL-21 and IL-4 in the lupus-like context. Splenocytes from the indicated mice were stimulated with PMA and Ionomycin (P/I +) or not (P/I–) as described in the methods section. Intracellular flow cytometry was realized to visualize the indicated cytokine productions on the indicated cellular compartments. (**a,d**) Representative contour plots showing PMA-Ionomycin-induced IFNγ, IL-21, and IL-4 productions by TFH cells (as defined in **Fig. 2c**) in splenocytes from aged *Mcpt8*^*DTR*^ or *Lyn*^*–/–*^ *Mcpt8*^*DTR*^ mice depleted in basophils (DT+) or not (DT–) (see **methods**). (**b,e,g**) Proportions (%) of IL-21 (**b**), IL-4 (**e**), and IFNγ (**g**) producing cells among TFH cells in splenocytes stimulated with PMA and Ionomycin (P/I +) or not (P/I–) from aged *Mcpt8*^*DTR*^ (blue bars) or *Lyn*^*–/–*^ *Mcpt8*^*DTR*^ (red bars) mice depleted in basophils (DT+, lighter color) or not (DT–, darker color). (**c,f,h**) Proportions (%) of IL-21 (**c**), IL-4 (**f**), and IFNγ (**h**) producing cells among TFH cells in splenocytes stimulated with PMA and Ionomycin (P/I +) or not (P/I–) from PBS-injected mice (blue bars) and pristane-injected *Mcpt8*^*DTR*^ mice (red bars), basophil sufficient (DT–, darker color) or basophil-depleted (DT+, lighter color). (**i**) Representative contour plots showing non-stimulated (NS) and PMA-Ionomycin-induced (P/I) IL-6 and IL-4 productions by spleen basophils from aged *Mcpt8*^*DTR*^ or *Lyn*^*–/–*^ *Mcpt8*^*DTR*^ mice. (**j,k**) Proportions (%) of IL-6 (**j**) and IL-4 (**k**) producing cells among basophils in splenocytes stimulated with PMA and Ionomycin (P/I +) or not (P/I–) from aged *Mcpt8*^*DTR*^ (blue bars) or *Lyn*^*–/–*^ *Mcpt8*^*DTR*^ (red bars) mice. (**l,m**) Proportions (%) of IL-6 (**l**) and IL-4 (**m**) producing cells among basophils in splenocytes stimulated with PMA and Ionomycin (P/I +) or not (P/I–) from PBS-injected *Mcpt8*^*DTR*^ (blue bars) or pristane-injected *Mcpt8*^*DTR*^ (red bars) mice. (**b,c, e**-**h** and **j**-**m**) Results are from at least three independent experiments and presented as individual values in bars representing the mean values. (**b,c, e**-**h**) Statistical analyses were done by one-way ANOVA followed by Holm-Šídák’s multiple comparisons test between the indicated groups. (**j**-**m**) Statistical analyses were done by Mann-Whitney U test between the indicated groups. NS: not significant, p>0.05; *: p<0.05; **: p<0.01; ***: p<0.001; ****: p<0.0001.

Both IL-6 and IL-4 are involved, respectively, in TFH and TFH2 cell differentiation^7,15^. Basophils are potent producers of these cytokines^20,21^. We next assessed whether basophils in both pristane-injected and Lyn-deficient animals were more prone to produce these cytokines with or without PMA-Ionomycin restimulation. Unlike what was observed in basophils from control mice, constitutive IL-4 and IL-6 productions were significantly detected in non-restimulated basophils from both lupus-like models, whereas no major differences were noticed after PMA-ionomycin restimulation (**Fig. 3i-m**). These results suggested that during the course of the disease, along with their increased expression of PD-L1 (**Fig. 2a,b**), basophils produce IL-6 and IL-4 *in vivo* which may explain some of their effects on the expansion of the TFH cell population and its TFH2 cell bias.

### Basophils promote *ex vivo* TFH cell differentiation through IL-4- and PD-L1-dependent mechanisms

Next, we evaluated the effects of basophils and basophil-expressed mediators on the differentiation of TFH cells *ex vivo* in a co-culture system. The presence of basophils induced a clear differentiation of the CD3/CD28-activated naïve CD4^+^ T cells towards the TFH cell subset with an increased ability of these TFH cells to produce IL-21 and IL-6 (**Fig. 4a-c**). Moreover, the basophil-induced TFH cell differentiation was biased towards the TFH2 cell subset as evidenced by an increased ability to produce IL-4 and IL-13 (**Fig. 4d,e**). We next bred CT-M8 (or *Mcpt8*^*CT/CT*^) mice^12^ with *Il4*^*fl/fl* 22^, *Il6*^*fl/fl* 23^, or *Pdl1*^*fl/fl* 24^ mice and generated mice deficient for IL-4, IL-6, or PD-L1 selectively in the basophil compartment (**Fig. S4a-c**). IL-4-deficient basophils could not induce any of the effects on the TFH cell differentiation as compared to WT basophils (**Fig. 4a-e**). IL-6-deficient basophils could still induce TFH cell differentiation despite reduced IL-6, but increased IL-21, production by TFH cells (**Fig. 4a-c**) and had a limited effect on the TFH2 cell differentiation (**Fig. 4d,e**). Interestingly, PD-L1 expression by basophils was mandatory to induce TFH cell differentiation but had a limited impact on the TFH2 cell differentiation that was completely dependent on basophil-expressed IL-4 (**Fig. 4a-e**). Of note, PMA-ionomycin restimulation of the co-cultured cells in the aforementioned conditions led to the same conclusions (**Fig. S4d-g**).

**Figure 4:**
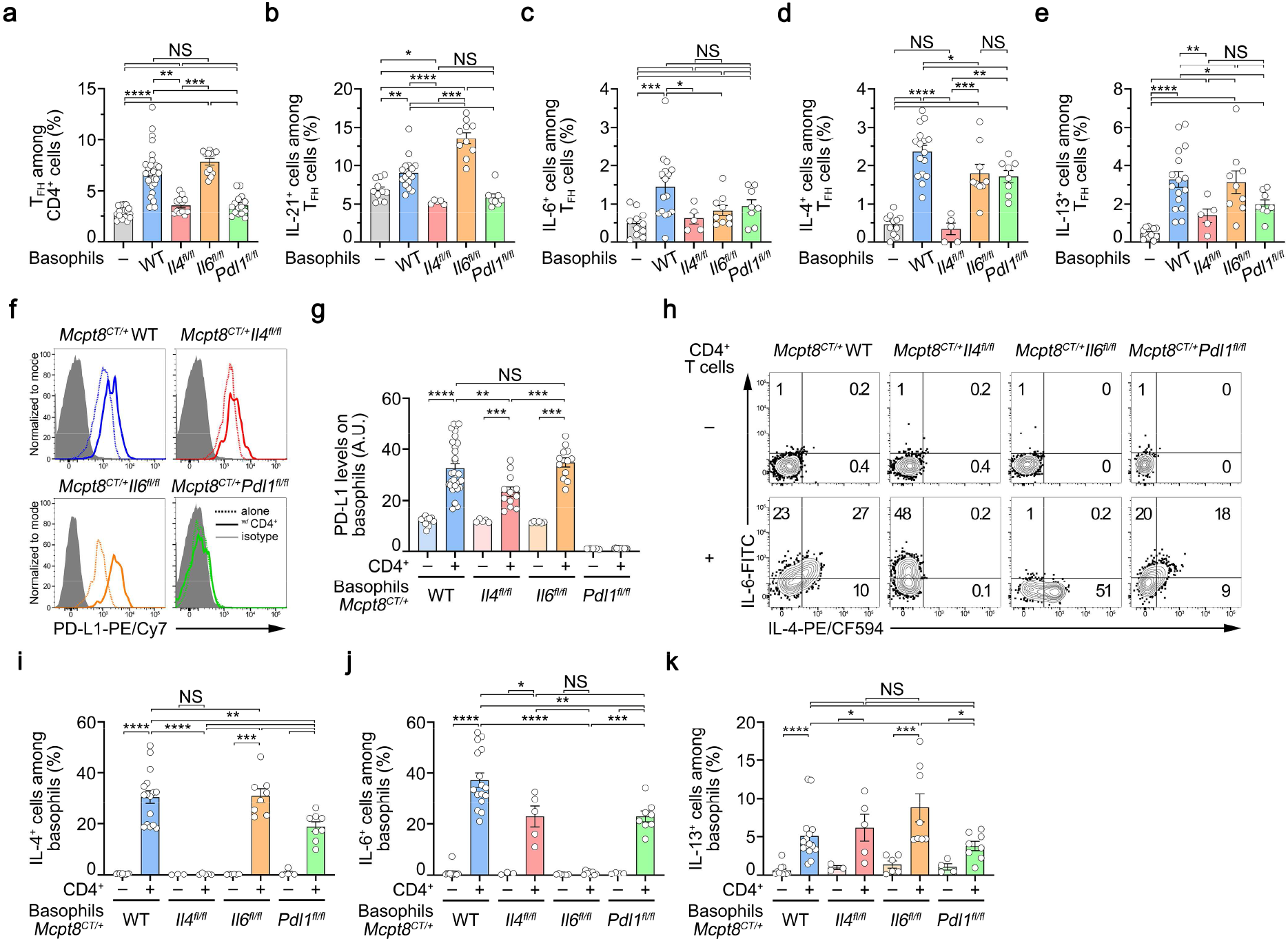
A bidirectional crosstalk between basophils and CD4^+^ T cells promotes *ex vivo* TFH differentiation. (**a**-**k**) Purified wild-type naïve CD4^+^ T cells were cultured in plates coated with anti-CD3 and anti-CD28 antibodies for three days without (–, grey filled bars) or with purified basophils from the following mice: *Mcpt8*^*CT/+*^ (WT) (blue filled bars), *Mcpt8*^*CT/+*^ *Il4*^*fl/fl*^ (*Il4*^*fl/fl*^, IL-4-deficient basophils) (red filled bars), *Mcpt8*^*CT/+*^ *Il6*^*fl/fl*^ (*Il6*^*fl/fl*^, IL-6-deficient basophils) (orange filled bars), or *Mcpt8*^*CT/+*^ *Pdl1*^*fl/fl*^ (*Pdl1*^*fl/fl*^, PD-L1-deficient basophils) (green filled bars). (**a**) Proportions (%) of TFH among CD4^+^ T cells after three days of culture as described in **methods**. (**b**-**e**) Proportions (%) of IL-21- (**b**), IL-6- (**c**), IL-4- (**d**), and IL-13- (**e**) producing cells among TFH cells non-restimulated and incubated with brefeldin A for the last 4 hours of the culture. (**f**) Representative histograms of PD-L1 expression on purified spleen basophils, from mice with the indicated genotype, co-cultured for three days without (dotted lines) or with activated naïve CD4 T cells (solid lines) (as described above). (**g**) PD-L1 expression levels on basophils, from mice as described above, cultured without (–, light colors) or with (+, dark colors) purified wild-type CD3/CD23-activated naïve CD4^+^ T cells for three days. (**h**) Representative contour plots showing IL-6 and IL-4 spontaneous production of basophils of the indicated genotype after three days of culture without (–) or with activated CD4+ T cells and in the presence of brefeldin A for the last 4 hours of culture. (**i,j,k**) Proportions (%) of spontaneous IL-4- (**i**), IL-6- (**j**), and IL-13- (**k**) producing cells among basophils of the indicated genotypes cultured as described above. (**a**-**k**) Results for the same cells but re-stimulated with PMA and ionomycin are shown in **Fig. S4d-j**. (**a**-**e, g, i**-**k**) Results are from at least three independent experiments and presented as individual values in bars representing the mean ± s.e.m.. (**a**) Statistical analyses were done by a Kruskal-Wallis test followed by Dunn’s multiple comparisons tests. (**b**-**e, g, i**-**k**) Statistical analyses were done by Mann-Whitney U test between the indicated groups. NS: not significant, p>0.05; *: p<0.05; **: p<0.01; ***: p<0.001; ****: p<0.0001.

Altogether, these results demonstrated that basophils induce *ex vivo* TFH cell differentiation in an IL-4- and PD-L1-dependent manner and that basophil-derived IL-6 is dispensable for these effects.

### CD4^+^ T cells promote basophil ability to induce TFH cell differentiation *ex vivo*

In parallel, we analyzed the effects of the CD4^+^ T cells on the basophil compartment. The presence of CD4^+^ T cells up-regulated PD-L1 expression on basophils and induced constitutive IL-4, IL-6, and IL-13 productions by these cells (**Fig. 4f-k**). The CD4^+^ T cell-induced up-regulation of PD-L1 on basophils was partially dependent on the IL-4 produced by basophils themselves, but independent of basophil-derived IL-6 (**Fig. 4f,g**). Conversely, the CD4^+^ T cell-induced IL-4 (and IL-6) production by basophils was partially dependent on PD-L1 expressed by basophils without impacting their maximal production ability (**Fig. 4h-i**, and **Fig. S4h,i**). IL-6 deficiency in basophils did not alter their ability to produce IL-4 constitutively in the presence of CD4^+^ T cells nor after PMA-ionomycin restimulation but enhanced their IL-13 production only in the former situation (**Fig. 4h-k** and **Fig. S4h-j**). These results suggest that PD-L1 engagement on basophils by CD4^+^ T cells, beyond promoting IL-21 production by TFH cells (**Fig. 4b**), was responsible for the extent of T cell-induced cytokine production by basophils. This may explain why PD-L1 deficient basophils could not promote TFH cell differentiation (**Fig. 4a**). Of note, CD4^+^ T cells induced IL-13 production by basophils and enhanced their maximal ability to produce this cytokine independently of their IL-4 or PD-L1 expression (**Fig. 4k** and **Fig. S4j**).

Altogether, these results strongly suggest that a bi-directional interaction between naïve CD4^+^ T cells and basophils leads to the differentiation of TFH cells via a mechanism mainly depending on IL-4 and PD-L1 expression by basophils but independent of the basophil-derived IL-6.

### PD-L1 controls the basophil-TFH cell functional relationship during lupus-like disease

We next sought to validate *in vivo* the relevance of PD-L1 expression by basophils in their functional relationship with TFH cells in the lupus-like context. Pristane injection did not lead to TFH cell accumulation in SLO in mice with basophil-restricted PD-L1-deficiency (*Mcpt8*^*CT/+*^ *Pdl1*^*fl/fl*^) as compared to WT animals (*Mcpt8*^*CT/+*^) (**Fig. 5a** and **Fig. S5**). However, PD-L1 expression by basophils was not required for basophil accumulation in SLO (**Fig. 5b**). Both the lupus-like context *in vivo* (**Fig. 3l,m**) and the co-culture system *in vitro* (**Fig. 4h-k**) induced IL-4 and IL-6 production by basophils. Here, PD-L1 deficiency on basophils completely prevented these cytokine basal productions in the lupus-like context (**Fig. 5c,d**). This was associated with a non-expansion of CD19^+^CD138^+^ cells in SLO (**Fig. 5e**) leading to dramatically reduced anti-RNP autoreactive IgG titers in the blood (**Fig. 5f**). Moreover, increased IgG and C3 glomerular deposits were barely detectable in the kidney of pristane-injected *Mcpt8*^*CT/+*^ *Pdl1*^*fl/fl*^ mice as compared to their WT counterparts (**Fig. 5g,h**).

**Figure 5:**
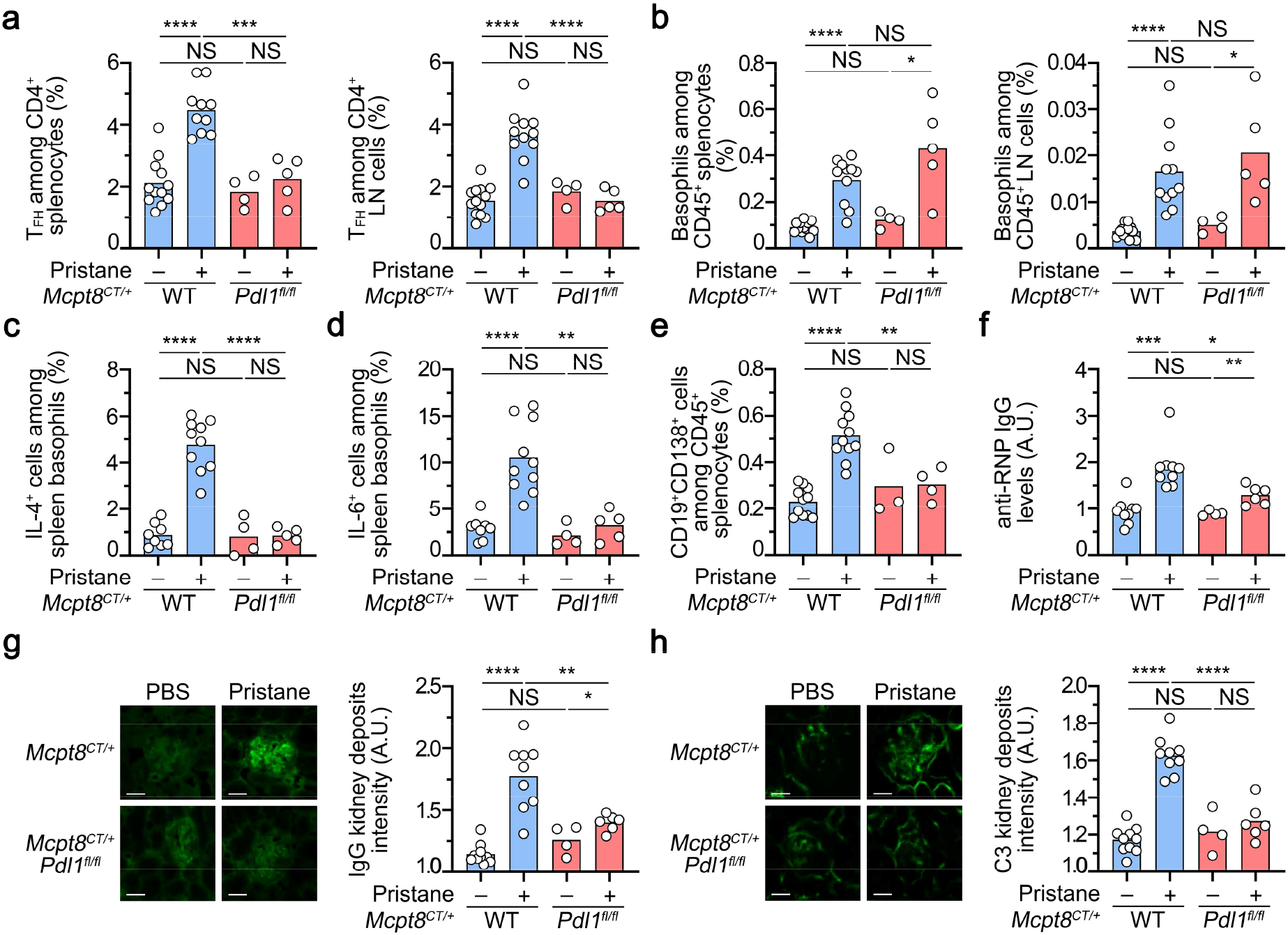
PD-L1 controls the basophil-TFH functional relationship during lupus-like disease. (**a**) Proportions (%) of TFH among CD4^+^ T cells in spleen (left) and lymph nodes (right) from *Mcpt8*^*CT/+*^ (WT) (blue filled bars) or *Mcpt8*^*CT/+*^ *Pdl1*^*fl/fl*^ (*Pdl1*^*fl/fl*^) (red filled bars) mice injected with PBS (–) or with pristane (+). (**b**) Proportions (%) of basophils among CD45^+^ cells in the spleen (left) and lymph nodes (right) from the mice described in (**a**). (**c,d**) Proportions (%) of spontaneous IL-4^+^ (**c**) or IL-6^+^ cells (**d**) among basophils in the spleen from the mice described in (**a**) incubated 4 hours in the presence of brefeldin A. (**e**) Proportions (%) of CD19^+^CD138^+^ cells among CD45^+^ cells in spleen from the mice described in (**a**). (**f**) Anti-RNP IgG autoantibody plasma titers from the same mice as in (**a**) were quantified by ELISA, as described in the methods section. O.D. values at 450 nm were determined and data were normalized to the mean of PBS-injected *Mcpt8*^*CT/+*^ values. (**g,h**) **Left**, Representative pictures of one glomerulus from mice with the indicated genotypes treated without (PBS, –) or with pristane (+) showing the intensity of anti-IgG (**g**) or anti-C3 (**h**) staining by immunofluorescence. Scale bar = 50 μm. Uncropped images are shown in **Fig. S6a** (IgG) and **S6b** (C3). **Right**, quantification of IgG (**g**) and C3 (**h**) glomerular deposits in kidneys from the mice described in (**a**). (**a**-**h**) Results are from at least three independent experiments and presented as individual values in bars representing the mean values. Statistical analyses were done by unpaired Student t-tests between the indicated groups. NS: not significant, p>0.05; *: p<0.05; **: p<0.01; ***: p<0.001; ****: p<0.0001.

Altogether, these results strongly suggest that PD-L1 basophil expression and up-regulation during lupus development were not involved in basophil accumulation in SLO but were responsible for the basophil-induced promotion of TFH and short-lived plasma cell expansions and subsequent pathological parameters.

### Basophil-derived IL-4 exerts a dual effect during lupus-like disease development

Since TFH cell differentiation *ex vivo* depended on IL-4 expression by basophils (**Fig. 4**), we next sought to verify whether basophil-derived IL-4 was mandatory for the basophil-TFH cell functional relationship in the lupus-like context. As suggested above, IL-4 deficiency selectively in the basophil compartment (*Mcpt8*^*CT/+*^ *Il4*^*fl/fl*^) prevented the pristane-induced TFH cell accumulation in SLO as compared to WT (*Mcpt8*^*CT/+*^) animals (**Fig. 6a**). By contrast, basophil recruitment into SLO upon pristane treatment of the mice was not dependent on basophil-derived IL-4 since basophil accumulation in these locations in mice was similar in *Mcpt8*^*CT/+*^ *Il4*^*fl/fl*^ and *Mcpt8*^*CT/+*^ WT animals (**Fig. 6b**). Surprisingly, CD19^+^CD138^+^ cells still accumulated in SLO despite selective basophil IL-4 deficiency (**Fig. 6c**). This phenotype was associated with a still significant constitutive IL-6 production by IL-4-deficient basophils in the lupus-like context (**Fig. 6d**). However, no significant titers of anti-RNP IgG autoantibodies were detected in the blood of pristane-injected *Mcpt8*^*CT/+*^ *Il4*^*fl/fl*^ mice as compared to their WT counterparts. In line with this feature, no IgG deposits were present in the glomeruli of pristane-treated mice with basophil-specific IL-4 deficiency but C3 deposits, although reduced, were still detected (**Fig. 6f** and **Fig. S6a,b**). Along with the accumulation of CD19^+^CD138^+^ cells in the SLO, the latter result led us to assess the presence of autoreactive IgM in the pristane-treated *Mcpt8*^*CT/+*^ *Il4*^*fl/fl*^ mice. As suspected, increased titers of anti-RNP IgM in the plasma of mice with IL-4 deficient basophils were observed as compared to pristane-treated *Mcpt8*^*CT/+*^ WT mice (**Fig. 6g**). These autoreactive IgM were not detected in the blood from pristane-injected basophil-specific PD-L1-deficient mice nor constitutive basophil-deficient mice (**Fig. 6g**) in line with the absence of plasmablast accumulation in SLO from these mice (**Fig. 5e** and ^12^). These anti-RNP IgM titers correlated with the presence of IgM deposits in the glomeruli of the corresponding mice (**Fig. 6h,i**). Altogether, these results strongly suggest that TFH cell pathogenic accumulation in SLO and autoreactive IgG titers induced by pristane were dependent on the basophil-derived IL-4. However, CD19^+^CD138^+^ short-lived plasma cell accumulation, which is dependent on basophils (**Fig. 2** and ^11,12,18^), was not dependent on IL-4 production by basophils. This led to an accumulation of plasmatic autoreactive anti-RNP IgM, with no IgG deposits but increased C3 deposits in glomeruli of the *Mcpt8*^*CT/+*^ *Il4*^*fl/fl*^ mice, suggesting that basophil-derived IL-4 was both enabling autoreactive antibody switch towards the IgG isotype and TFH cell accumulation that promoted this phenomenon as well^7,15^.

**Figure 6:**
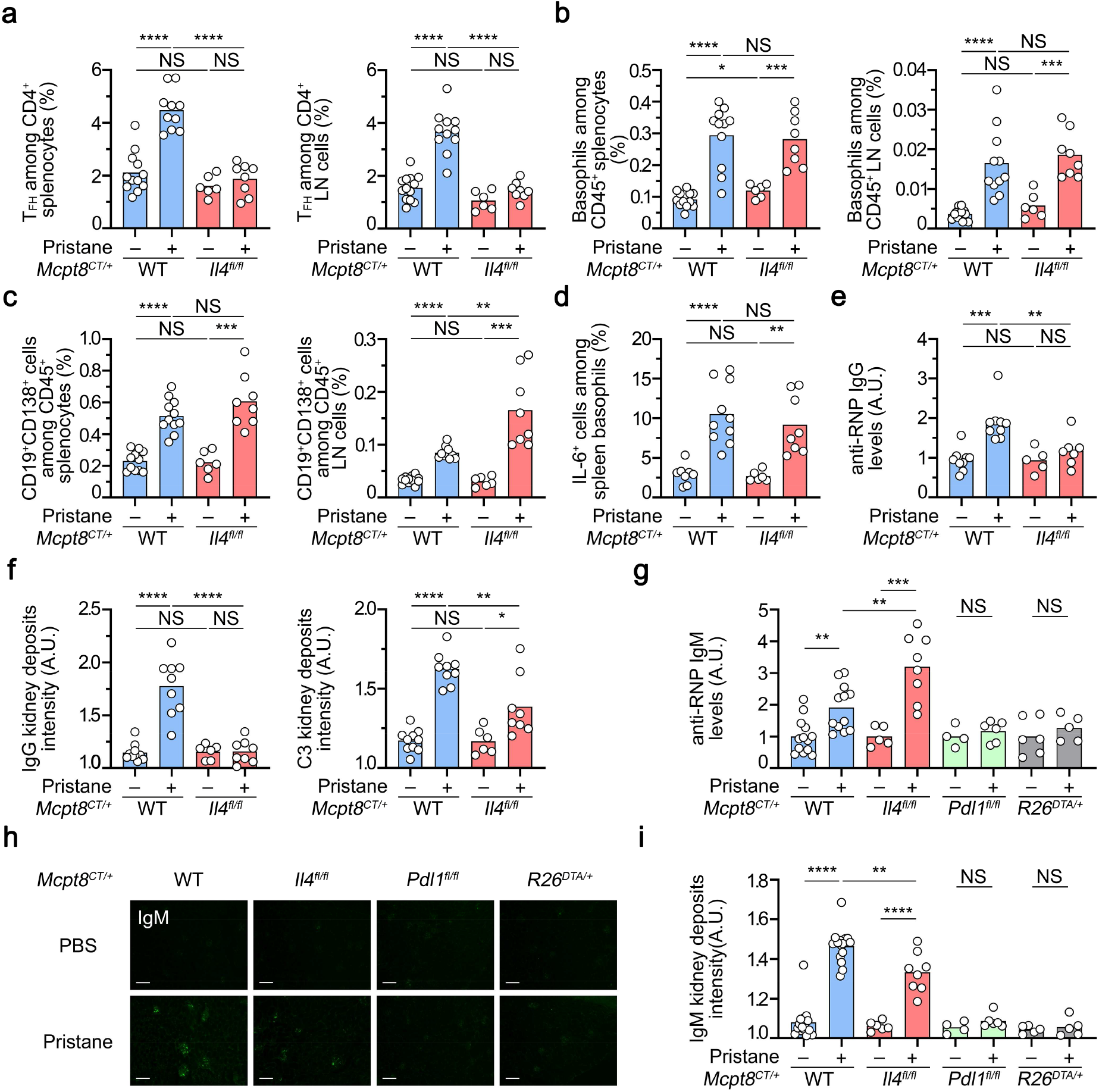
Basophil-derived IL-4 controls T-dependent autoreactive antibody isotype switch in lupus-like disease. (**a**) Proportions (%) of TFH among CD4^+^ T cells in spleen (left) and lymph nodes (right) from *Mcpt8*^*CT/+*^ (WT) (blue filled bars) or *Mcpt8*^*CT/+*^ *Il4*^*fl/fl*^ (*Il4*^*fl/fl*^) (red filled bars) mice injected with PBS (–) or with pristane (+). (**b**) Proportions (%) of basophils among CD45^+^ cells in the spleen (left) and lymph nodes (right) from the mice described in (**a**). (**c**) Proportions (%) of CD19^+^CD138^+^ cells among CD45^+^ cells the in the spleen (left) and lymph nodes (right) from the mice described in (**a**). (**d**) Proportions (%) of spontaneous IL-6^+^ cells among basophils in the spleen from the mice described in (**a**) incubated 4 hours in the presence of brefeldin A. (**e**) Anti-RNP IgG autoantibody plasma levels from the same mice as in (**a**) were quantified by ELISA, as described in the methods section. O.D. values at 450 nm were determined and data were normalized to the mean of PBS-injected *Mcpt8*^*CT/+*^ values. (**f**) Quantification of C3 (left) and IgG (right) glomerular deposits in kidneys from the mice described in (**a**). A representative picture for each genotype in each condition is shown in **Fig. S6a,b**. (**a**-**f**) The data shown that concerns *Mcpt8*^*CT/+*^ (WT) mice are the same as the data shown in **Fig. 5**. (**g**) Anti-RNP IgM autoantibody plasma levels were determined by ELISA, as described in the methods section. O.D. values at 450 nm were determined and data were normalized to the mean of the values of PBS-injected control mice for each genotype. The mice analyzed were *Mcpt8*^*CT/+*^ (WT), *Mcpt8*^*CT/+*^ *Il4*^*fl/fl*^ (*Il4*^*fl/fl*^), *Mcpt8*^*CT/+*^ *Pdl1*^*fl/fl*^ (*Pdl1*^*fl/fl*^) (green filled bars) and basophil-deficient (*Mcpt8*^*CT/+*^ *R26*^*DTA/+*^) (*R26*^*DTA/+*^, grey filled bars) mice treated with PBS or pristane (pristane – or +, respectively). (**h**) Representative pictures of kidneys from mice with the indicated genotypes injected with PBS or pristane showing the intensity of anti-IgM staining by immunofluorescence. Scale bar = 200 μm. (**i**) Quantification of IgM glomerular deposits in kidneys from the mice described in (**g**). (**a**-**i**) Results are from at least three independent experiments and presented as individual values in bars representing the mean values. (**a**-**g,i**) Statistical analyses were done by unpaired Student t-tests between the indicated groups. NS: not significant, p>0.05; *: p<0.05; **: p<0.01; ***: p<0.001; ****: p<0.0001.

### Human basophils drive *ex vivo* TFH cell and TFH2 cell differentiation through IL-4, IL-6, and PD-1-dependent mechanisms

We next sought to validate the ability of basophils to promote TFH cell differentiation in a human co-culture system. First, CD3- and CD28-activated human naïve CD4^+^ T cells were cultured for three days without or with increasing numbers of purified human basophils demonstrating the capacity of human blood basophils to induce TFH cell differentiation (**Fig. 7a-c)** more potently than mouse spleen basophils (**Fig. 4a**). Next, cultures of CD3- and CD28-activated human naïve CD4^+^ T cells alone or together with basophils were repeated in the presence of blocking antibodies targeting IL-4, IL-6, PD-1, or their corresponding isotype controls. As seen in the murine system, IL-4 and PD-1 antagonisms led to a dramatic decrease in the ability of human basophils to drive CD4^+^ T cell differentiation into TFH cells (**Fig. 7d**). IL-6 blockade also decreased basophil-induced TFH cell differentiation, probably due to its effects on the T cell-derived IL-6 (**Fig. 7d**). Most of the basophil-induced TFH cells were belonging to the TFH2 cell subset expressing neither CCR6 nor CXCR3 (**Fig. 7e,f**). This TFH2 cell subset differentiation was dramatically dampened by the blockade of IL-4, IL-6, or PD-1 (**Fig. 7e,f**).

**Figure 7:**
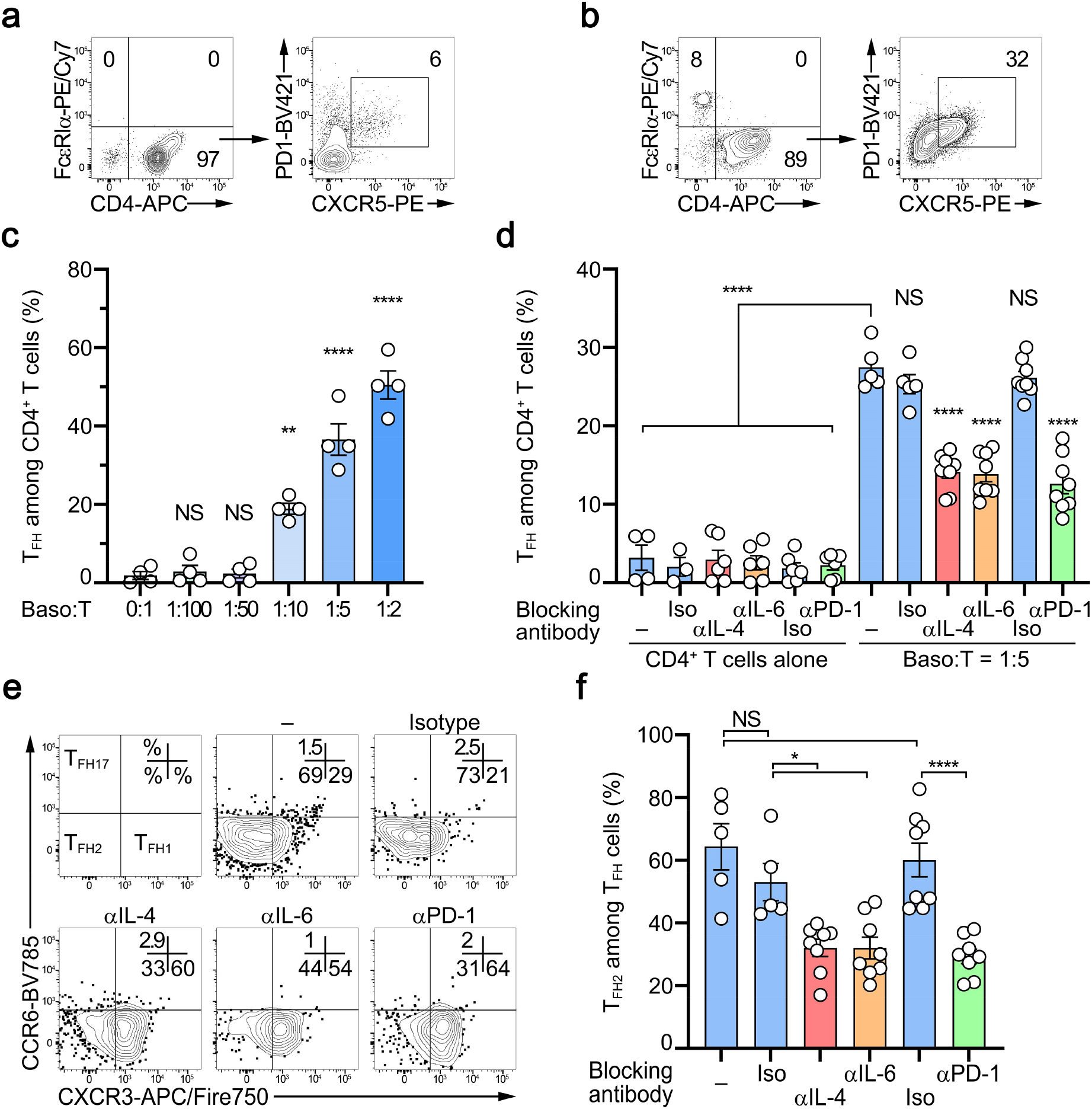
Human basophils drive TFH differentiation through IL-4, IL-6, and PD-L1 dependent mechanisms *ex vivo*. (**a,b**) Representative contour plots showing CD3/CD28-activated human CD4^+^ T cells cultured for three days without (**a**) or with (**b**) purified human basophils at a 5:1 ratio (left panels) and the condition-induced TFH differentiation of the CD4+ T cells (right panels). Basophils were defined as FcεRIα^+^ CRTH2^+^ CCR3^+^ cells and TFH cells were defined as CD4^+^ PD1^+^ CXCR5^+^ ICOS^+^ cells. (**c**) Proportions (%) of TFH among CD3/CD28-activated CD4^+^ T cells after three days of culture without (0:1) or with the indicated ratio of purified human basophils. (**d**) Proportions (%) of TFH cells among CD3/CD28-activated CD4^+^ T cells after three days of culture without (CD4^+^ T cells alone) or with purified human basophils at a ratio of 1:5 in the absence (–) (blue filled bars) or presence of antibodies blocking IL-4 (αIL-4) (red filled bars), IL-6 (αIL-6) (orange filled bars) or PD-1 (αPD-1) (green filled bars) or the corresponding isotype controls (Iso) (blue filled bars). (**e**) Representative contour plots showing subsets of TFH cells as defined in the upper left panel based on CCR6 and CXCR3 expressions on cells as in (**a**). (**f**) Proportions (%) of TFH2 cells among TFH cells after culture as described in (**d**). TFH2 cells were defined as CD4^+^ PD1^+^ CXCR5^+^ ICOS^+^ CCR6^−^ CXCR3^−^ cells. (**c,d**) Data are presented as individual values in bars representing the mean values ± s.e.m. (**c**) One representative experiment with cells from 4 different donors is shown. (**d,f**) Results are from three independent experiments. (**c,d,f**) Statistical analyses were done by one-way ANOVA test followed by Tukey’s multiple comparisons tests between the indicated groups. NS: not significant, p>0.05; **: p<0.01; ***: p<0.001; ****: p<0.0001.

Altogether, these data demonstrate that human basophils could induce human naïve CD4^+^ T cell differentiation into TFH cells, especially into the TFH2 cell subset, and that this was dependent on IL-4 and PD-1. Together with the PD-L1 overexpression by blood basophils from SLE patients (**Fig. 1e**), these results strongly suggest that, as demonstrated in the lupus-like mouse models (**Figs. 2-6)**, basophils promote the pathogenic accumulation of TFH cells during SLE pathogenesis through their expression of PD-L1 and IL-4.

## DISCUSSION

In this study, we have identified mechanisms by which basophils control the pathogenic accumulation of TFH cells in SLO to promote autoreactive IgG production during SLE pathogenesis.

In a normal antigen-driven immunization process, germinal centers (GC) are key structures that allow B cell maturation into high affinity and class-switched antibody-secreting cells^25^. The formation and maintenance of these structures depend on TFH cells^7^. Basophils are dispensable to mount an efficient humoral response to OVA protein immunization^19^ and do not control basal TFH or OVA-induced TFH cell populations (this study). Importantly, in the latter conditions, basophils do not accumulate in SLO in contrast to the lupus-like context. Dysregulated expansion of TFH cells in an SLE-like context occurs through spontaneous GC-like reactions which are favored by the abnormal cytokine milieu and mainly in the extrafollicular (EF) area^7^. Our results indicate that the pathogenic accumulation of TFH cells in SLO in a lupus-like context, and thus spontaneous TFH cell responses, are fully dependent on SLO-localized basophils and more precisely mediated by basophil expression of PD-L1 and IL-4.

The impairment of this basophil-dependent pathogenic accumulation of TFH cell in SLO led to a dramatic reduction in autoreactive anti-RNP IgG production and IgG kidney deposits, further validating the relevance of both cell types in autoantibody and pathogenic CIC productions in a lupus-like context. Investigating whether basophils control spontaneous EF TFH cell responses in other autoimmune diseases such as rheumatoid arthritis or multiple sclerosis may lead to developing common therapeutic strategies for these different autoimmune conditions that may share some pathophysiological pathways^26^.

Recently, Kim *et al*. showed that TFH2 cells are induced by, and produce some, IL-4. These TFH2 cells are central in humoral autoimmunity, promote autoreactive IgE production with their frequencies increased in both SLE patients and also some SLE-like mouse models^15^. Accordingly, systemic IL-4 blockade dampened TFH2 cell accumulation and their deleterious effects in the *Ets1*^*ΔCD4*^ lupus-like mouse model. As exogenous IL-4 induces Gata3 expression in T cells enabling TH2 cell differentiation^27^, exogenous IL-4 enabled Gata3^+^ TFH2 cell accumulation in an SLE-like context in a similar way^15^. Our data strongly suggest that basophils deliver the required IL-4-induced priming to CD4^+^ T cells in a PD-L1-dependent manner that allows TFH and TFH2 cell accumulation in the lupus-like context. Further studies will be required to validate this feature in other lupus-like mouse models including *Ets1*^*ΔCD4*^ mice. This IL-4 mediated basophil-TFH pathogenic axis is important since basophil depletion may represent a safer therapeutic strategy than global IL-4 neutralization. Indeed, this cytokine controls a large number of physiologically protective processes^28,29^ and other deleterious pathways involved in SLE pathophysiology may limit the benefits for the patients of long-term IL-4 blockade^2^.

Both human and murine basophils promoted CD3/CD28 activated naïve CD4^+^ T cell differentiation into TFH cells *ex vivo* in the absence of specific antigen or recognized potent antigen-presenting cells. These effects depended on basophil-expressed IL-4 and PD-L1 (but not on IL-6) for murine cells and were dampened by IL-4, IL-6, or PD-1 blocking antibodies for human cells. This setting may resume spontaneous TFH cell responses occurring in the SLE context^7^. TCR repertoire analysis of these basophil-induced TFH cells may help to decipher whether these spontaneous TFH cell responses favor autoreactivity of the TFH cell compartment.

pDC and type I IFN have recently been shown to promote EF T-dependent B cell responses to extracellular self DNA^30^. Our data suggest that basophils and their PD-L1 expression are mandatory to induce TFH cell pathogenic accumulation and B cell response to nuclear antigens. PD-L1 expression by CD11c-expressing cells inhibits TFH cell response in experimental autoimmune encephalomyelitis^31^. Type I IFN promotes human basophil apoptosis, but this effect is rescued by IL-3^32^ whose production (mainly by T cells^33^) is increased during SLE pathogenesis^34,35^ and acts as well on pDC survival^36^. Thus, it may be relevant to study the interplay between basophils and DC subsets in this process. pDC or follicular DC and basophils may indeed compete to engage PD-1 on TFH cells to respectively inhibit or promote TFH cell expansion and TFH2 cell differentiation in EF responses in an autoimmune context.

PD-L1 expression by basophils is known in humans and may be induced by IFNγ^37^. PD-L1 upregulation on human basophils has recently been described during SARS-CoV2 infections^38^ although it is not directly induced by the virus^39^. Interestingly, basopenia occurs in acute SARS-CoV2 infection suggesting putative recruitment of basophils to SLO^40^. This hypothesis is further supported by basophil overexpression of CXCR4 and CD62L in the SARS-CoV-2 infection setting, similar to what we described in SLE patients^10,11,40,41^. Normalization of both blood basophil numbers and activation markers is associated with the humoral response and recovery of COVID-19 patients^40,42^. This may indicate a role for PD-L1-overexpressing basophils in TFH cell induction to promote antiviral humoral response. EF responses occurring in lupus-prone mice and during some infections can quickly involute if new plasmablasts are not generated^25^. Thus, controlling basophil function or localization in SLO during viral infections may represent interesting therapeutic strategies to promote or sustain the antiviral humoral response.

Unexpectedly, when injected with pristane, mice with IL-4-deficient basophils still showed some plasmablast accumulation in SLO and C3 kidney deposits despite the absence of TFH cell accumulation, anti-RNP IgG induction, and IgG kidney deposits. Unlike pristane-injected basophil-deficient mice or mice with PD-L1-deficient basophils, *Mcpt8*^*CT/+*^*Il4*^*fl/fl*^ mice developed high titers of anti-RNP IgM. This indicates that basophils express factors other than IL-4 in SLO during lupus-like disease responsible for their PD-L1-dependent action on B cell maturation into plasmablast in the absence of TFH cell pathogenic accumulation. However, it strongly suggests as well that basophil-derived IL-4 is mandatory for autoantibody class switch towards IgG in the lupus-like context. Further characterization of this B cell–basophil relationship in SLO in the SLE context will be required to identify basophil-derived factors that contribute to TFH-independent plasmablast proliferation. Our study establishes a direct link between basophils and TFH cells in the SLE context that promotes autoreactive IgG production and lupus nephritis pathogenesis. Altering the basophil/TFH cell axis in the SLE context may represent a promising innovative intervention strategy in SLE.

## Data Availability

All data produced in the present study are available upon reasonable request to the authors

## Acknowledgments

This work was supported by the Fondation pour la Recherche Médicale (FRM) [grant # EQU201903007794] to N.C., the French Agence Nationale de la Recherche (ANR) [grants # ANR-19-CE17-0029 BALUMET to N.C. and ANRPIA-10-LABX-0017 INFLAMEX], the Ministerio de Economía y Competitividad y Fondo Europeo de Desarrollo Regional (RTI2018-101105-B-I00) to J.H., by the Centre National de la Recherche Scientifique (CNRS), by Université Paris-Cité and by the Institut National de la Santé et de la Recherche Médicale (INSERM). We acknowledge the expert work from the members of the animal core facility (I. Renault and S. Olivré), the flow cytometry core facility (G. Gautier, J. Da Silva and V. Gratio) and the imaging facility (S. Benadda) of the Centre de Recherche sur l’Inflammation (INSERM UMR1149), and the help from O. Thibaudeau and L. Wingertsmann from the morphology core facility (INSERM UMR1152).

## Authors’ contributions

J.T. designed experiments, conducted experiments, and wrote the manuscript. N.C. conceived the project, designed experiments, conducted experiments, wrote the manuscript, and directed the project. Q.S., L.C., Y.L., F.S., E.P., J.B-C, C.P., U.B., M.B., G.H., K.S., and E.D. conducted experiments, analyzed the data, and/or edited the manuscript. H.K. and K.M. provided the *Il4*^*fl/fl*^ and *Mcpt8*^*DTR*^ mice and edited the manuscript. J.H. provided the *Il6*^*fl/fl*^ mice and edited the manuscript. P.G.F. provided the *Pdl1*^*fl/fl*^ mice and edited the manuscript. N.C. had full access to all of the data in the study and take responsibility for the integrity of the data and the accuracy of the data analysis. All authors approved the final version of the article.

## MATERIAL AND METHODS

### Patients

Blood samples were collected from adult patients enrolled in a prospective long-term study of systemic lupus erythematosus (SLE) and chronic renal diseases. All SLE patients fulfilled the American College of Rheumatology (ACR) classification criteria for SLE. SLE and healthy control (CT) donor characteristics are shown in **Table S1**. SELENA-SLEDAI (Safety of Estrogens in Lupus Erythematosus National Assessment - SLE Disease Activity Index) scores were assessed to evaluate patients’ lupus activity who were classified as inactive (0-1), mild (2-4), and active (> 4). The study was approved by the Comité Régional de Protection des Personnes (CRPP, Paris, France) under the reference ID-RCB 2014-A00809-38. SLE samples were obtained from in- and outpatients and clinical data were harvested after approval by the Commission Nationale de l’Informatique et des Libertés (CNIL). All samples were collected in heparinized tubes (BD vacutainer) and processed within 4 hours. Written informed consent was obtained from all individuals.

### Mice

*Mcpt8*^*DTR* 43^, *Lyn*^*—/—*^ *Mcpt8*^*DTR* 11,44^, *Il4*^*fl/fl* 22^, *Il6*^*fl/fl* 23^,and *Pdl1*^*fl/fl* 24^ mice were on a pure C57BL/6J background and bred in our animal facilities. Rosa26-loxP-Stop-loxP-DTA C57BL/6J (*R*_*26*_ ^*DTA/DTA*^ or *R*_*26*_ ^*DTA/+*^) mice^45^ were purchased from The Jackson Laboratory through Charles River Laboratories. CT-M8 (*Mcpt8*^*tm1*.*1(cre)lcs*^ or *Mcpt8*^*CT/CT*^ or *Mcpt8*^*CT/+*^) mice were recently described^12^. The mice crossed in our animal facilities (*Mcpt8*^*CT/+*^; *Mcpt8*^*CT/+*^ *Il4*^*fl/fl*^; *Mcpt8*^*CT/+*^ *Il6*^*fl/fl*^; *Mcpt8*^*CT/+*^ *Pdl1*^*fl/fl*^ and *Mcpt8*^*CT/+*^ *R*_*26*_ *DTA/+* were on a C57BL6J/N mixed genetic background at the F2 generation. For lupus-like disease analysis of the *Lyn*^*—/—*^ model, “aged” *Mcpt8*^*DTR*^ and *Lyn*^*—/—*^ *Mcpt8*^*DTR*^ age-matched and sex-matched mice were analyzed between 30 and 45 weeks of age (50% males and 50 % females). Mice were maintained under specific pathogen-free conditions in our animal facilities. The study was conducted in accordance with the French and European guidelines and approved by the local ethics committee comité d’éthique Paris Nord N°121 and the Ministère de l’enseignement supérieur, de la recherche et de l’innovation under the authorization number APAFIS#14115.

### Human samples handling

Heparinized human blood samples were centrifuged at 600 g for 5 minutes and 2 mL of plasma were collected and stored at -80°C for later analysis. Red blood cells (RBC) were lysed in RBC lysing buffer (150 mM NH_4_Cl, 12 mM NaHCO_3_, 1 mM EDTA, pH 7.4) in a ratio of 5mL of blood for 20mL of ACK lysing buffer. After 5 min of incubation at room temperature (rt), 25mL of PBS were added and cells were centrifuged at 600 g for 5 min, and the supernatant was discarded. This procedure was repeated 3 times. Leukocytes were then resuspended in fluorescence-activated cell sorting (FACS) buffer (PBS 1% BSA, 0.01% NaN_3_, 1mM EDTA) and prepared for flow cytometry (see below). Leukocyte count and viability (>95%) were assessed by trypan-blue staining on a Malassez hemacytometer.

### Pristane-induced lupus-like mouse model

Pristane-induced lupus-like disease was initiated by injecting 500 μL of 2,6,10,14-tetramethylpentadecane or Pristane (Sigma) into the peritoneal cavity of 7-10 weeks-old female mice. For control individuals, genotype- and age-matched female mice were injected with 500 μL of phosphate buffer saline pH 7.4 (PBS) (Gibco). Mice were attributed to PBS- or pristane-injected groups randomly and maintained in the same cage for the whole procedure. Pristane- and PBS-injected mice were analyzed 8 weeks after injection.

### DT-mediated basophil depletion in lupus-like context

For diphtheria toxin (DT)-mediated basophil depletion, *Mcpt8*^*DTR*^, and *Lyn*^*—/—*^ *Mcpt8*^*DTR*^ mice were injected intraperitoneally with 100 μL of PBS containing (or not) 1 μg of DT (Sigma) 10, 9, 6, 2 and 1 day before the sacrifice.

### Mouse OVA immunization experiments

10 to 15 weeks old C57/BL6J *Mcpt8*^*DTR*^ sex-matched mice were immunized by intraperitoneal injection of 200 μL of a 50/50 emulsion of Alum (Thermofisher Scientific) with 100 μg of ovalbumin (OVA, Sigma-Aldrich) diluted in PBS or with PBS alone for control mice. A similar injection was performed on day 7 and the mice were sacrificed on day 14. For DT-mediated basophil depletion, 1 μg of DT (or PBS as a control) was injected intraperitoneally on day 12 and day 13.

### Mouse samples processing

Mice were euthanized in a controlled CO_2_ chamber (TEM Sega) and blood sampling was performed through cardiac puncture with a heparin-coated syringe with a 25G needle. Blood was centrifuged at 300 g for 15 min and plasma was harvested and kept at -80°C for later analysis. RBC were lysed in 5mL of RBC lysing buffer for 5 min at rt and washed with 10mL of PBS. This procedure was repeated 3 times and cells were resuspended in PBS. The left kidney was harvested and embedded in OCT embedding matrix (Cellpath) and snap-frozen in liquid nitrogen and kept at -80°C for later analysis.

Spleen and lymph nodes (cervical, brachial and inguinal) were harvested in PBS and dissociated by mechanical disruption on a 40 μm cell strainer (Falcon, Corning). For splenocytes, RBC were lysed once in 5mL RBC lysing buffer 5 min at rt and washed with 10mL of PBS. Cell counts were assessed by trypan-blue staining on a Malassez hemacytometer and 1 to 5 million cells were used per FACS staining condition.

### *Ex vivo* stimulation of splenocytes

Mouse splenocytes were harvested as described above and resuspended at 5 million cells/mL in culture medium (RPMI 1640 with Glutamax and 20mM HEPES, 1mM Na-pyruvate, non-essential amino acids 1X (all from Life Technologies), 100 μg/mL streptomycin and 100 U/mL penicillin (GE Healthcare) and 37.5 μM β-mercaptoethanol (Sigma-Aldrich) supplemented with 20% heat-inactivated fetal calf serum (FCS) (Life Technologies)). For phorbol-myristate-acetate (PMA) and ionomycin stimulation experiments, whole splenocytes were stimulated or not with 40 nM of PMA and 800 nM ionomycin for 4 hours in the presence of 2 μg/mL of brefeldin A (all from Sigma Aldrich, Merck) and cultured at 37°C and 5% CO2. Then, cells were harvested by repeated flushing, and wells were washed with 1mL of PBS. Samples were then prepared for flow cytometry analysis.

### Flow cytometry staining

For human leukocytes, non-specific antibody binding sites were saturated with 20 μL of a solution containing 100 μg/mL of human, mouse, rat, and goat IgG (Jackson ImmunoResearch Europe and Innovative Research Inc.) in FACS buffer. 200 μL of staining solution containing the panel of fluorophore-conjugated specific antibodies or their fluorophore-conjugated isotypes (described in the **antibodies and reagents table**) were added to the cells for 30 min at 4°C protected from light. After a wash in PBS, cells were fixed in fixation buffer (Biolegend) for 20 minutes at 4°C and then washed in FACS buffer before data acquisition. For mouse samples, cells washed in PBS were stained with GHOST 510 viability dye (TONBO) following the manufacturer’s instructions. Non-specific antibody binding sites were saturated with 10 μg/mL of anti-CD16/CD32 antibody clone 2.4G2 (BioXCell), and 100 μg/mL of polyclonal rat, mouse, and Armenian Hamster IgG (Innovative Research Inc.) in FACS buffer and stained with the antibodies described in the **antibodies and reagents table** for 30 min in the dark at 4°C. Cells were then washed in FACS Buffer before data acquisition. For intracellular staining, cells were first stained extracellularly as described above. Cells were washed in PBS and fixed with fixation buffer for 20 min at 4°C. Cell permeabilization and intracellular staining were realized with permeabilization/wash buffer (Biolegend) following the manufacturer’s instructions. Cells were then resuspended in FACS buffer before acquisition All flow cytometry acquisitions were realized using a Becton Dickinson 5 lasers LSR Fortessa X-20 and data analysis using Flowjo vX (Treestar, BD Biosciences). For assessment of surface marker expression levels, ratios of the geometric mean fluorescence intensity (gMFI) of the markers to the gMFI of the corresponding isotype control were calculated and expressed in arbitrary units (A.U.).

### Kidney Immunofluorescence assays

4 μm thick cryosections of OCT-embedded kidneys were fixed 20 min in ice-cold acetone and kept a - 80°C until immunofluorescence staining. Slides were thawed and fixed in 10% formalin (Sigma) for 20 min at room temperature and blocked with PBS containing 1% BSA (Euromedex) and 5% goat serum (Sigma-Aldrich). Slices were stained for 2 hours at room temperature in the dark in a humid chamber with FITC-conjugated anti-mouse C3 (Cedarlane), Alexa Fluor® 488-conjugated anti-mouse C3 (Santa Cruz Biotech), Alexa Fluor® 488-conjugated goat anti-mouse IgG Fcγ-specific (Jackson Immunoresearch), FITC-conjugated goat anti-mouse IgM (BioRad (AbD Serotec)) or corresponding isotype controls. Slides were mounted in Immunomount (Thermofischer Scientifics) and analyzed by fluorescent microscopy (Leica DMR, Leica microsystems). The ratio of specific glomerular fluorescence over tubulointerstitial background was then measured using ImageJ software v. 1.43u (NIH), averaging at least 30 glomeruli per mouse for each sample.

### Anti-RNP IgG and IgM autoantibody detection

Maxisorp 96 well plates (Thermo Scientific) were coated overnight at 4°C with 10 μg/mL of purified RNP complexes (Immunovision) diluted in carbonate buffer (100 mM NaHCO_3_ and 30 mM Na_2_CO_3_ pH 9.6). Plates were washed 3 times in PBS containing 0.1% of Tween-20 (Bio-Rad laboratories) (PBS-T) and saturated for 1 hour with PBS containing 5% of FCS. For anti-RNP IgG quantification, plasma samples were diluted 1:25 in PBS-T containing 5% of goat serum and 100 μL added to the wells.

Samples, positive and negative controls were incubated for 2 hours at rt. Plates were washed 5 times with PBS-T. 100 μL of either 500 ng/mL goat anti-mouse IgG (Invitrogen) or 10 ng/mL goat anti-mouse IgM (Bethyl laboratories) conjugated to horseradish peroxidase (HRP) diluted in PBS-T containing 5% goat serum were added and incubated 1h at room temperature. After 5 washes in PBS-T, 100 μL of tetramethylbenzidine (TMB) substrate (ThermoFisher) were added to the wells and incubated at least 20 min at rt and the reaction was stopped with 0.2N sulfuric acid solution. Optical density at 450 nm was measured by spectrophotometry (Infinite 200 Pro plate reader, TECAN). On each plate, similar negative and positive controls were run. Optical density (OD) values were first normalized to the negative controls and then, the presented results were normalized to the mean of the control mice values and expressed in arbitrary units (A.U.).

### Anti-dsDNA IgG detection

Maxisorp 96 well plates (Thermo Scientific) were coated overnight at 4°C with calf thymus dsDNA (Sigma-Aldrich) diluted in TE buffer (Tris-HCl 10 mM, EDTA 1 mM pH 9) at a concentration of 2 μg/mL and diluted in the same volume of Pierce DNA coating solution (Thermo Scientific) to obtain a final concentration of 1μg/mL of dsDNA. The same protocol as for anti-RNP IgG was then followed but the PBS-T contained 0.05% of Tween-20.

### Cell sorting and co-culture

For mouse co-culture experiments, F(ab’)2 anti-CD3 (clone145-2C11 at 0,5 μg/mL) and anti-CD28 (clone PV-1 at 0,5 μg/mL) antibodies (BioXcell) were coated overnight at 4°C in culture 96 well plates (Costar) in sterile-filtered coating buffer (20 mM carbonate buffer (pH 9.6) containing 2 mM MgCl_2_ and 0.01% NaN_3_). Wells were washed in PBS before plating the cells. Spleens were harvested and handled as described above and resuspended at 10^8^ cells/mL in PBS containing 2% FCS and 2mM EDTA. Naïve CD4^+^ T cells were sorted by magnetic negative selection following manufacturer protocol (Stemcell Technologies) and resuspended in culture medium at 0.5 million cells/mL. 100 μL of naïve CD4^+^ T cell suspension were then added to wells of interest. Basophils from CTM8 mice were purified over 98% by electronic sorting using the BD FACSMelody Cell Sorter (BD Biosciences) using the tandem tomato fluorescent protein as a specific marker^12^. Basophils were re-suspended in culture medium at 0.1 million cells/mL and 100μL were added to wells of interest. All wells were supplemented with 10 pg/mL of recombinant mouse IL-3 (Peprotech). After 3 days of culture at 37°C 5% CO2, cells were harvested and prepared for flow cytometry as described above. For cytokine production detection, cells were stimulated or not for the last 4 hours with 40 nM PMA and 800 nM ionomycin in the presence of 2 μg/mL brefeldin A (Sigma-Aldrich, Merck) and prepared for flow cytometry as described above.

For human co-culture experiments, 96 well plates (Costar) were coated overnight at 4°C in coating buffer containing 5 μg/mL of mouse anti-human CD3 (clone OKT3) (Thermo Fischer Scientific). Blood from healthy volunteers was handled and lysed as described above in sterile conditions and re-suspended at 5×10^7^ cells/mL in PBS 2% FCS (Gibco) 2mM EDTA. Naïve CD4+ T cells and basophils were sorted by magnetic negative selection following manufacturer instructions (Stemcell Technologies). Naïve CD4^+^ T cells were re-suspended in culture medium at 0.5 million cells/mL. and 100 μL of cell suspension were then added to each well. Basophils were resuspended in culture medium at 0.1 million cells/mL and 100μL were added to each well unless otherwise specified. All wells were supplemented with 10 pg/ml of recombinant human IL-3 (Biolegend) and 2 μg/mL of mouse anti-human CD28 antibody (Clone CD28.2 at (BioXcell)). When indicated, some wells were supplemented with mouse anti-human blocking antibodies targeting IL4 (Clone MP4-25D2), IL-6 (Clone MQ2-13A5), both at 5 μg/mL or PD-1 (Clone EH12.2H7 at 10μg/mL) or with the corresponding isotypes at the same concentrations. Cells were harvested after 3 days of culture at 37°C 5% CO_2_ and prepared for flow cytometry as described above.

### Statistical analysis

Two-tailed Student’s unpaired t-tests were used to compare the differences of one variable between two groups when distributions were Gaussian and Mann-Whitney U tests for non-parametric distributions. When more than two groups were compared, one-way ANOVA coupled with *ad hoc* multiple comparisons post-tests, as indicated in the figure legends depending on the distribution of the values in each group, were used. Statistical analyses were realized using Prism v9.4 software (Graphpad).

### Antibodies, reagents, software and equipments

See Supplementary Table S3.

## GRAPHICAL ABSTRACT

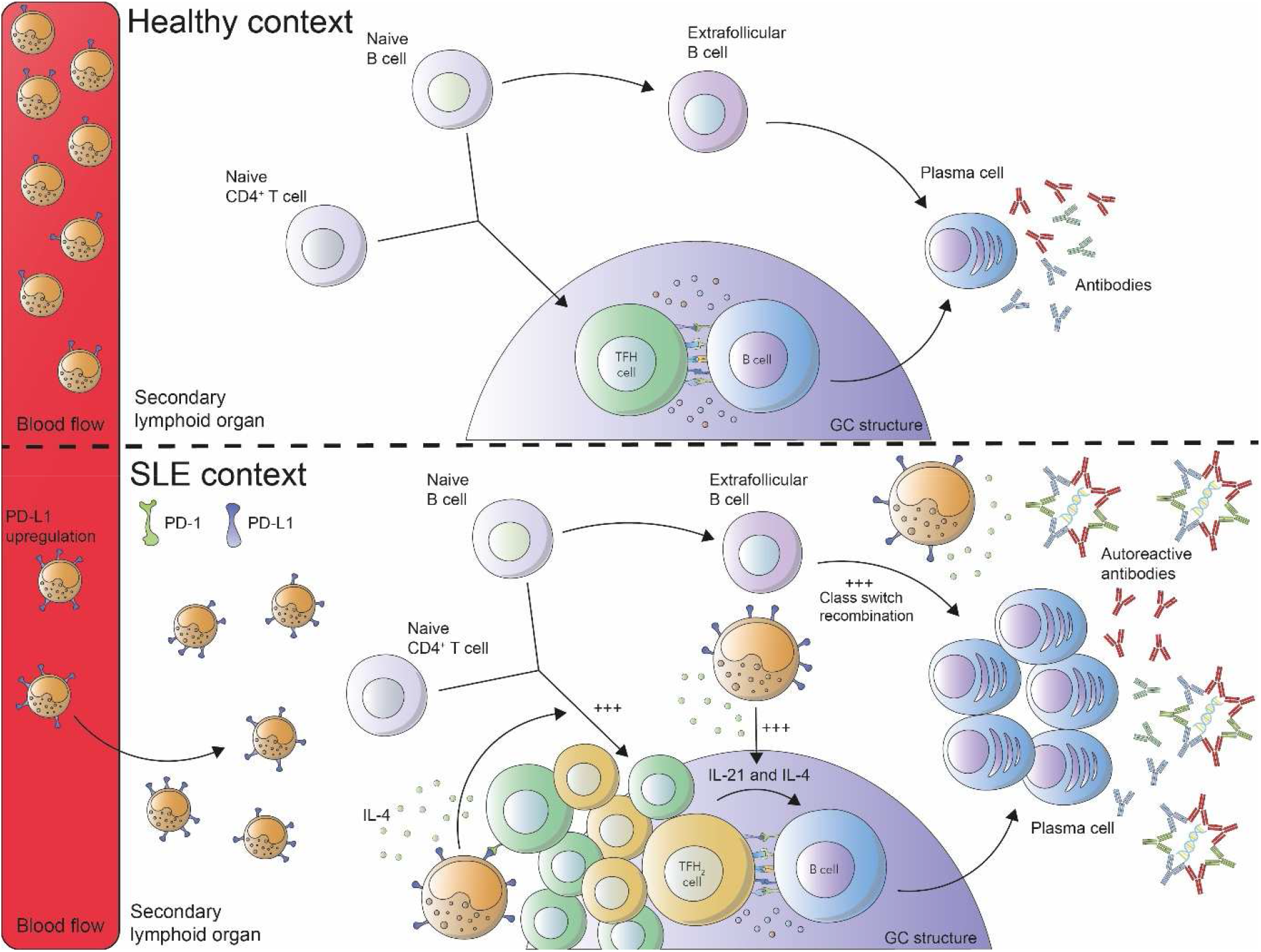

Pathogenic accumulation of T follicular helper cells in lupus disease depends on PD-L1 and IL-4 expressing basophils.

