## Supplemental Information for "Pathogenic accumulation of T follicular helper cells in lupus disease depends on PD-L1 and IL-4 expressing basophils"

**SUPPLEMENTARY INFORMATION**

**Authors**: John TCHEN [1,2], Quentin SIMON [1,2], Léa CHAPART [1,2], Yasmine LAMRI [1,2], Fanny SAIDOUNE [1,2], Emeline PACREAU [1,2], Christophe PELLEFIGUES [1,2], Julie BEX-COUDRAT [1,2], Hajime KARASUYAMA [3], Kensuke MIYAKE [3], Juan HIDALGO [4], Padraic G. FALLON [5], Thomas PAPO [1,2,7], Ulrich BLANK [1,2], Marc BENHAMOU [1,2], Guillaume HANOUNA [1,2,6], Karim SACRE [1,2,7], Eric DAUGAS [1,2,6] and Nicolas CHARLES [1,2]

Affiliations:

1 Université Paris Cité, Centre de Recherche sur l'Inflammation, INSERM UMR1149, CNRS EMR8252, Faculté de Médecine site Bichat, Paris, France

2 Université Paris Cité, Laboratoire d'Excellence Inflamex, Paris, France

3- Inflammation, Infection and Immunity Laboratory, TMDU Advanced Research Institute, Tokyo Medical and Dental University (TMDU), Tokyo, Japan

4- Universidad Autonoma de Barcelona, Facultad de Biociencias, Unidad de Fisiologia Animal Bellaterra, Barcelona, Spain

5- School of Medicine, Trinity College Dublin, Dublin, Ireland.

6- Service de Néphrologie, Hôpital Bichat, Assistance Publique – Hôpitaux de Paris, Paris, France

7- Service de Médecine Interne, Hôpital Bichat, Assistance Publique – Hôpitaux de Paris, Paris, France

*__Correspondence to__:

Nicolas Charles, PhD

Centre de Recherche sur l'Inflammation, INSERM UMR1149, CNRS ERL8252,

Université Paris Cité, Faculté de Médecine site Bichat,

16 rue Henri Huchard, 75018 Paris, France.

ORCID : **0000-0002-5416-5834**

**Supplementary Table S1. SLE Patient and healthy control characteristics**

| Variables | SLE patients | | | | Healthy controls |
|---|---|---|---|---|---|
| | ALL SLE | Inactive (SLEDAI≤1) | Mild (1<SLEDAI≤4) | Active (SLEDAI>4) | |
| ***Demographic characteristics*** | | | | | |
| n | 204 | 54 | 51 | 99 | 43 |
| Age, mean±SD, yr | 38.87±12.4 | 44.43±13.5 | 39.4±11.9 | 35.6±10.9 | 34±9.2 |
| Female, n (%) | 184 (90) | 47 (87) | 47 (92) | 90 (90) | 32 (74) |
| ***Lupus characteristics*** | | | | | |
| Disease duration, mean±SD, yr | 9.5±8.5 | 14.2±9.0 | 10.4±7.3 | 7.00±7.8 | - |
| Anti-dsDNA Ab positive, n (%) | 111 (54) | 7 (12) | 26 (51) | 78 (79) | - |
| History of lupus nephritis, n (%) | 163 (80) | 32 (59) | 37 (73) | 89 (90) | - |
| SLEDAI | | | | | |
| Mean±SD | 6.2±6.3 | 0.0±0.2 | 3.2±0.9 | 11.2±5.5 | - |
| Median (Min-Max) | 4 (0-33) | 0 (0-1) | 4 (2-4) | 10 (5-33) | - |
| ***Treatment characteristics*** | | | | | |
| Current prednisone dose (mg/day) | | | | | |
| Mean±SD | 13.5±38.8 | 3.7±3.8 | 7.8±7.9 | 22.2±54.8 | - |
| 15mg/day or higher, n (%) | 39 (19) | 1 (2) | 8 (16) | 30 (30) | - |
| Concurrent immuno-suppressive therapy (n, %) | | | | | |
| hydroxychloroquine | 174 (85) | 43 (80) | 46 (90) | 82 (83) | - |
| mycophenolate mofetil | 67 (33) | 18 (33) | 22 (43) | 27 (27) | - |
| IV cyclophosphamide | 13 (6) | 1 (2) | 1 (2) | 11 (11) | - |
| azathioprine | 16 (8) | 0 (0) | 5 (10) | 11 (11) | - |

SD: standard deviation; yr: year; IV: intravenous.

**Supplementary Table S2.**

**Expression levels of some T cell interacting surface molecules on basophils from healthy controls and SLE patients**

| | Healthy control basophils | | | SLE patient basophils | | | |
|---|---|---|---|---|---|---|---|
| Surface Marker | Mean | SD | n | Mean | SD | n | p-value |
| PD-L1 | 5.603 | 2.028 | 39 | 8.455 | 4.308 | 204 | <0.0001 |
| PD-L2 | 0.9547 | 0.1361 | 5 | 1.085 | 0.2912 | 42 | 0.2737 |
| ICOSL | 1.676 | 0.2078 | 5 | 1.757 | 0.5207 | 56 | 0.9492 |
| OX40L | 2.499 | 1.842 | 18 | 2.647 | 1.92 | 178 | 0.8469 |
| CD84 | 21.41 | 4.981 | 20 | 26.09 | 11.2 | 74 | 0.0956 |

Mean corresponds to the ratio of the geometric mean fluorescence intensity (gMFI) of the indicated surface marker and the gMFI of the isotype control as determined by flow cytometry. SD: standard deviation. Statistical analyses were done by Mann-Whitney U tests.

43 **Supplementary Table S3.**

| REAGENT | SOURCE | IDENTIFIER |
|---|---|---|
| **Antibodies** | | |
| Alexa Fluor 488 anti-mouse Complement Component C3 (Clone: 11H9) | Santa Cruz Biotechnology | (Santa Cruz Biotechnology Cat# sc-58926, RRID:AB_1119819) |
| Alexa Fluor 488 anti-mouse/human CD44 (Clone: IM7) | Biolegend | ( Cat# 103016, RRID:AB_493679) |
| Alexa Fluor 488 anti-mouse IgG Fcγ fragment specific (Goat Polyclonal) | Jackson ImmunoResearch | (Jackson ImmunoResearch Labs Cat# 115-545-008, RRID:AB_2338842) |
| Alexa Fluor 647 anti-mouse FcεRIα (Clone: MAR-1) | Biolegend | ( Cat# 134310, RRID:AB_1626093) |
| Alexa Fluor 647 anti-mouse CD3ε (Clone: 145-2C11) | Biolegend | ( Cat# 100322, RRID:AB_389322) |
| Alexa Fluor 700 anti-mouse CD45 (Clone: 30-F11) | Biolegend | ( Cat# 103128, RRID:AB_493715) |
| Alexa Fluor 700 anti-mouse CD62L (Clone: MEL-14) | Biolegend | ( Cat# 104426, RRID:AB_493719) |
| APC/Fire 750 anti-mouse CD4 (Clone: RM4-5) | Biolegend | ( Cat# 100568, RRID:AB_2629699) |
| APC/Cyanine7 anti-mouse CD4 (Clone: RM4-5) | Biolegend | ( Cat# 100526, RRID:AB_312727) |
| APC/Cyanine7 anti-mouse TCR β chain (Clone: H57-597) | Biolegend | ( Cat# 109220, RRID:AB_893624) |
| APC/Fire 750 anti-mouse TCR β chain (Clone: H57-597) | Biolegend | ( Cat# 109246, RRID:AB_2629697) |
| APC/Cyanine7 anti-mouse CD19 (Clone: 6D5) | Biolegend | ( Cat# 115530, RRID:AB_830707) |
| APC/Fire 750 anti-mouse CD19 (Clone: 6D5) | Biolegend | ( Cat# 115558, RRID:AB_2572120) |
| APC/Cyanine7 anti-mouse CD117 (c-kit) (Clone: 2B8) | Biolegend | ( Cat# 105826, RRID:AB_1626278) |
| APC/Fire 750 anti-mouse CD117 (c-Kit) (Clone: 2B8) | Biolegend | ( Cat# 105838, RRID:AB_2616739) |
| Biotin anti-mouse CD185 (CXCR5) (Clone: L138D7) | Biolegend | ( Cat# 145510, RRID:AB_2562126) |
| Biotin anti-mouse CD4 (Clone: RM4-5) | Biolegend | ( Cat# 100508, RRID:AB_312711) |
| Biotin anti-mouse CD19 (Clone: 6D5) | Biolegend | ( Cat# 115504, RRID:AB_313639) |
| Biotin anti-mouse CD8a (Clone: 53-6.7) | Biolegend | ( Cat# 100704, RRID:AB_312743) |
| Biotin anti-mouse NK-1.1 (Clone: PK136) | Biolegend | ( Cat# 108704, RRID:AB_313391) |
| BUV395 anti-mouse CD45 (Clone: 30-F11) | BD Biosciences | (BD Biosciences Cat# 564279, RRID:AB_2651134) |
| Brilliant Violet 421 anti-mouse CD4 (Clone: GK1.5) | Biolegend | ( Cat# 100438, RRID:AB_11203718) |
| Brilliant Violet 421 anti-mouse CD279 (PD-1) (Clone: 29F.1A12) | Biolegend | ( Cat# 135218, RRID:AB_2561447) |
| Brilliant Violet 421 anti-mouse CD138 (Syndecan-1) (Clone: 281-2) | Biolegend | ( Cat# 142508, RRID:AB_11203544) |
| Brilliant Violet 421 anti-mouse/human CD44 (Clone: IM7) | Biolegend | ( Cat# 103040, RRID:AB_2616903) |
| Brilliant Violet 605 anti-mouse CD45 (Clone: 30-F11) | Biolegend | ( Cat# 103140, RRID:AB_2562342) |
| Brilliant Violet 605 anti-mouse CD279 (PD-1) (Clone: 29F.1A12) | Biolegend | ( Cat# 135220, RRID:AB_2562616) |
| Brilliant Violet 605 anti-mouse CD152 (Clone: UC10-4B9) | Biolegend | ( Cat# 106323, RRID:AB_2566467) |
| Brilliant Violet 785 anti-mouse CD19 (Clone: 6D5) | Biolegend | ( Cat# 115543, RRID:AB_11218994) |
| Brilliant Violet 785 anti-mouse/human CD44 (Clone: IM7) | Biolegend | ( Cat# 103059, RRID:AB_2571953) |

| | | |
|---|---|---|
| Brilliant Violet 785 anti-mouse CD274 (B7-H1, PD-L1) (Clone: 10F.9G2) | Biolegend | ( Cat# 124331, RRID:AB_2629659) |
| eFluor 450 anti-mouse IL-6 (Clone: MP5-20F3) | Thermo Fisher Scientific | (Thermo Fisher Scientific Cat# 48-7061-82, RRID:AB_2574103) |
| eFluor 450 anti-mouse IL-21 (Clone: FFA21) | Thermo Fisher Scientific | (Thermo Fisher Scientific Cat# 48-7211-82, RRID:AB_2811832) |
| FITC anti-mouse Complement Component C3 (Clone: RmC11H9) | Cedarlane | (CEDARLANE Cat# CL7503F, RRID:AB_10061294) |
| FITC anti-mouse IgM (Goat polyclonal) | BioRad (AbD Serotec) | (Bio-Rad Cat# 102002, RRID:AB_619870) |
| FITC anti-mouse CD123 (Clone: 5B11) | Thermo Fisher Scientific | (Thermo Fisher Scientific Cat# 11-1231-82, RRID:AB_465192) |
| FITC anti-mouse CD49b (Clone: HMα2) | Biolegend | ( Cat# 103504, RRID:AB_313027) |
| FITC anti-mouse IL-6 (Clone: MP5-20F3) | Thermo Fisher Scientific | (Thermo Fisher Scientific Cat# 11-7061-82, RRID:AB_465394) |
| FITC anti-mouse TNF-α (Clone: MP6-XT22) | R&D Systems | (R and D Systems Cat# IC410F, RRID:AB_357323) |
| Pacific Blue anti-mouse CD49b (pan-NK cells) (Clone: DX5) | Biolegend | ( Cat# 108918, RRID:AB_2265144) |
| PE anti-mouse IL-21 (Clone: FFA21) | Thermo Fisher Scientific | (Thermo Fisher Scientific Cat# 12-7211-82, RRID:AB_1834466) |
| PE anti-mouse CD278 (ICOS) (Clone: 15F9) | Biolegend | ( Cat# 107706, RRID:AB_313335) |
| PE anti-mouse FOXP3 (Clone: FJK-16s) | Thermo Fisher Scientific | (Thermo Fisher Scientific Cat# 12-5773-82, RRID:AB_465936) |
| PE-CF594 anti-mouse IL-4 (Clone: 11B11) | BD Biosciences | (BD Biosciences Cat# 562450, RRID:AB_2737616) |
| PE/Cyanine7 anti-mouse CD45 (Clone: 30-F11) | Biolegend | ( Cat# 103114, RRID:AB_312979) |
| PE/Cyanine7 anti-mouse IL-13 (Clone: eBio13A) | Thermo Fisher Scientific | (Thermo Fisher Scientific Cat# 25-7133-82, RRID:AB_2573530) |
| PE/Cyanine7 anti-mouse CD62L (Clone: MEL-14) | Biolegend | ( Cat# 104418, RRID:AB_313103) |
| PE/Cyanine7 anti-mouse CD274 (B7-H1, PD-L1) (Clone: 10F.9G2) | Biolegend | ( Cat# 124314, RRID:AB_10643573) |
| PE/Dazzle 594 anti-mouse CD279 (PD-1) (Clone: 29F.1A12) | Biolegend | ( Cat# 135228, RRID:AB_2566006) |
| PE/Dazzle 594 anti-mouse CD19 (Clone: 6D5) | Biolegend | ( Cat# 115554, RRID:AB_2564001) |
| PerCP/Cyanine5.5 anti-mouse CD8a (Clone: 53-6.7) | Biolegend | ( Cat# 100734, RRID:AB_2075238) |
| PerCP/Cyanine5.5 anti-mouse CD4 (Clone: RM4-5) | Biolegend | ( Cat# 100540, RRID:AB_893326) |
| PerCP/Cyanine5.5 anti-mouse IFN-γ (Clone: XMG1.2) | Biolegend | ( Cat# 505822, RRID:AB_961359) |
| Alexa Fluor 647 anti-human CD294 (CRTH2) (Clone: BM16) | Biolegend | ( Cat# 350104, RRID:AB_10642025) |
| Alexa Fluor 647 anti-human CD273 (B7-DC, PD-L2) (Clone: MIH18) | Biolegend | ( Cat# 345514, RRID:AB_2728313) |
| Alexa Fluor 700 anti-human/mouse/rat CD278 (ICOS) (Clone: C398.4A) | Biolegend | ( Cat# 313528, RRID:AB_2566126) |
| APC anti-human CD4 (Clone: RPA-T4) | Biolegend | ( Cat# 300537, RRID:AB_2562051) |
| APC anti-human CD275 (B7-H2, ICOSL) (Clone: 2D3) | Biolegend | ( Cat# 309408, RRID:AB_2565557) |
| APC/Cyanine7 anti-human CD183 (CXCR3) (Clone: G025H7) | Biolegend | ( Cat# 353722, RRID:AB_2561423) |
| Brilliant Violet 421 anti-human CD203c (E-NPP3) (Clone: NP4D6) | Biolegend | ( Cat# 324612, RRID:AB_2563848) |
| Brilliant Violet 421 anti-human CD274 (B7-H1, PD-L1) (Clone: 29E.2A3) | Biolegend | ( Cat# 329714, RRID:AB_2563852) |
| Brilliant Violet 421 anti-human CD279 (PD-1) (Clone: NAT105) | Biolegend | ( Cat# 367422, RRID:AB_2721517) |

| | | |
|---|---|---|
| Brilliant Violet 605 anti-human CD193 (CCR3) (Clone: 5E8) | Biolegend | ( Cat# 310716, RRID:AB_2563831) |
| Brilliant Violet 785 anti-human CD197 (CCR7) (Clone: G043H7) | Biolegend | ( Cat# 353230, RRID:AB_2563630) |
| BUV395 anti-human CD3 (Clone: SK7 (also known as Leu-4)) | BD Biosciences | (BD Biosciences Cat# 564000, RRID:AB_2744382) |
| BUV395 anti-human CD14 (Clone: MφP9 (also known as MφP-9)) | BD Biosciences | (BD Biosciences Cat# 563561, RRID:AB_2744288) |
| BUV395 anti-human CD56 (Clone: NCAM16.2 (also known as NCAM 16)) | BD Biosciences | (BD Biosciences Cat# 563554, RRID:AB_2687886) |
| BUV395 anti-human CD19 (Clone: SJ25C1 (also known as SJ25-C1)) | BD Biosciences | (BD Biosciences Cat# 563551, RRID:AB_2738274) |
| PE anti-human CD185 (CXCR5) (Clone: J252D4) | Biolegend | ( Cat# 356904, RRID:AB_2561813) |
| PE anti-human CD84 (Clone: CD84.1.21) | Biolegend | ( Cat# 326008, RRID:AB_2229003) |
| PE anti-human CD252 (OX40L) (Clone: 11C3.1) | Biolegend | ( Cat# 326308, RRID:AB_2207271) |
| PE/Cyanine7 anti-human FcεRIα (Clone: AER-37 (CRA-1)) | Biolegend | ( Cat# 334620, RRID:AB_10575314) |
| PE/Dazzle 594 anti-human CD123 (Clone: 6H6) | Biolegend | ( Cat# 306034, RRID:AB_2566450) |
| Alexa Fluor 488 Rat IgG2a, κ Isotype Control (Clone: RTK2758) | Biolegend | ( Cat# 400525, RRID:AB_2864283) |
| Alexa Fluor 488 Goat IgG whole molecule (Goat polyclonal) | Jackson ImmunoResearch | (Jackson ImmunoResearch Labs Cat# 005-540-003, RRID:AB_2337013) |
| Alexa Fluor 488 Rat IgG2b, κ Isotype Ctrl (Clone: RTK4530) | Biolegend | ( Cat# 400625, RRID:AB_389321) |
| Alexa Fluor 647 Armenian Hamster IgG Isotype Control (Clone: HTK888) | Biolegend | ( Cat# 400924, RRID:AB_2922967) |
| Alexa Fluor 647 Rat IgG2a, κ Isotype Control (Clone: RTK2758) | Biolegend | ( Cat# 400526, RRID:AB_2864284) |
| Alexa Fluor 647 Mouse IgG1, κ Isotype Control (Clone: MOPC-21) | Biolegend | ( Cat# 400130, RRID:AB_2800436) |
| Alexa Fluor 700 Rat IgG2a, κ Isotype Control (Clone: RTK2758) | Biolegend | ( Cat# 400528, RRID:AB_2923249) |
| Alexa Fluor 700 Mouse IgG1, κ Isotype Control (Clone: MOPC-21) | Biolegend | ( Cat# 400143, RRID:AB_2923250) |
| APC Mouse IgG1, κ Isotype Control (Clone: MOPC-21) | Biolegend | ( Cat# 400119, RRID:AB_2888687) |
| APC Mouse IgG2b, κ Isotype Control (Clone: MPC-11) | Biolegend | ( Cat# 400322, RRID:AB_326500) |
| APC/Cyanine7 Armenian Hamster IgG Isotype Control (Clone: HTK888) | Biolegend | ( Cat# 400927, RRID:AB_2923251) |
| APC/Cyanine7 Rat IgG2a, κ Isotype Control (Clone: RTK2758) | Biolegend | ( Cat# 400523, RRID:AB_2923252) |
| APC/Cyanine7 Rat IgG2b, κ Isotype Control (Clone: RTK4530) | Biolegend | ( Cat# 400623, RRID:AB_326565) |
| APC/Cyanine7 Mouse IgG1, κ Isotype Control (Clone: MOPC-21) | Biolegend | ( Cat# 400127, RRID:AB_2892538) |
| APC/Fire 750 Armenian Hamster IgG Isotype Control (Clone: HTK888) | Biolegend | ( Cat# 400961, RRID:AB_2923253) |
| APC/Fire 750 Rat IgG2a, κ Isotype Control (Clone: RTK2758) | Biolegend | ( Cat# 400567, RRID:AB_2923254) |
| APC/Fire 750 Rat IgG2b, κ Isotype Control (Clone: RTK4530) | Biolegend | ( Cat# 400669, RRID:AB_2905475) |
| Biotin Rat IgG2b, κ Isotype Control (Clone: RTK4530) | Biolegend | ( Cat# 400603, RRID:AB_326547) |
| BUV395 Rat IgG2b, κ Isotype Control (Clone: R35-38) | BD Biosciences | (BD Biosciences Cat# 563560, RRID:AB_2869507) |
| BUV395 Mouse IgG1, k Isotype Control (Clone: X-40) | BD Biosciences | (BD Biosciences Cat# 563547, RRID:AB_2869503) |
| BUV395 Mouse IgG2b, κ Isotype Control (Clone: 27-35) | BD Biosciences | (BD Biosciences Cat# 563558, RRID:AB_2869505) |
| Brilliant Violet 421 Rat IgG2a, κ Isotype Control (Clone: RTK2758) | Biolegend | ( Cat# 400535, RRID:AB_10933427) |

| | | |
|---|---|---|
| Brilliant Violet 421™ Rat IgG2b, κ Isotype Control (Clone: RTK4530) | Biolegend | ( Cat# 400639, RRID:AB_10895758) |
| Brilliant Violet 421 Mouse IgG1, κ Isotype Control (Clone: MOPC-21) | Biolegend | ( Cat# 400157, RRID:AB_10897939) |
| Brilliant Violet 421 Mouse IgG2b, κ Isotype Control (Clone: MPC-11) | Biolegend | ( Cat# 400341, RRID:AB_10898160) |
| Brilliant Violet 605 Armenian Hamster IgG Isotype Control (Clone: HTK888) | Biolegend | ( Cat# 400943, RRID:AB_2923255) |
| Brilliant Violet 605 Rat IgG2a, κ Isotype Control (Clone: RTK2758) | Biolegend | ( Cat# 400539, RRID:AB_11126979) |
| Brilliant Violet 605 Rat IgG2b, κ Isotype Control (Clone : RTK4530) | Biolegend | ( Cat# 400649, RRID:AB_2864282) |
| Brilliant Violet 785 Rat IgG2a, κ Isotype Control (Clone: RTK2758) | Biolegend | ( Cat# 400545, RRID:AB_11218993) |
| Brilliant Violet 785 Rat IgG2b, κ Isotype Control (Clone: RTK4530) | Biolegend | ( Cat# 400647, RRID:AB_2923256) |
| Brilliant Violet 785 Mouse IgG2a, κ Isotype Control (Clone: MOPC-173) | Biolegend | ( Cat# 400273, RRID:AB_2923257) |
| eFluor 450 Rat IgG1, κ Isotype Control (Clone: eBRG1) | Thermo Fisher Scientific | (Thermo Fisher Scientific Cat# 48-4301-82, RRID:AB_1271984) |
| eFluor 450 Rat IgG2a, κ Isotype Control (Clone: eBRG1) | Thermo Fisher Scientific | (Thermo Fisher Scientific Cat# 48-4321-82, RRID:AB_1271999) |
| FITC Armenian Hamster IgG Isotype Control (Clone: HTK888 | Biolegend | ( Cat# 400905, RRID:AB_2923258) |
| FITC Rat IgG1, κ Isotype Control (Clone: RTK2071) | Biolegend | ( Cat# 400405, RRID:AB_326511) |
| FITC Rat IgG2a, κ Isotype Control (Clone: RTK2758) | Biolegend | ( Cat# 400506, RRID:AB_2736919) |
| Pacific Blue Rat IgM, κ Isotype Control (Clone: RTK2118) | Biolegend | ( Cat# 400816, RRID:AB_10644001) |
| PE Mouse IgG1, κ Isotype Control (Clone: MOPC-21) | Biolegend | ( Cat# 400112, RRID:AB_2847829) |
| PE Mouse IgG2a, κ Isotype Ctrl Antibody (Clone: MOPC-173) | Biolegend | ( Cat# 400212, RRID:AB_326460) |
| PE-CF594 Rat IgG1, κ Isotype Control (Clone: R3-34) | BD Biosciences | (BD Biosciences Cat# 562309, RRID:AB_11153318) |
| PE/Cyanine7 Rat IgG1, κ Isotype Control (Clone: RTK2071) | Biolegend | ( Cat# 400416, RRID:AB_326522) |
| PE/Cyanine7 Rat IgG2a, κ Isotype Control (Clone: RTK275) | Biolegend | ( Cat# 400522, RRID:AB_326542) |
| PE/Cyanine7 Rat IgG2b, κ Isotype Control (Clone: RTK4530) | Biolegend | ( Cat# 400618, RRID:AB_326560) |
| PE/Cyanine7 Mouse IgG2b, κ Isotype Control (Clone: MPC-11) | Biolegend | ( Cat# 400325, RRID:AB_2923259) |
| PE/Dazzle 594 Rat IgG2a, κ Isotype Control (Clone: RTK2758) | Biolegend | ( Cat# 400557, RRID:AB_2923260) |
| PE/Dazzle 594 Mouse IgG1, κ Isotype Control (Clone: MOPC-21) | Biolegend | ( Cat# 400175, RRID:AB_2923261) |
| PerCP/Cyanine5.5 Rat IgG2a, κ Isotype Control (Clone: RTK2758) | Biolegend | ( Cat# 400531, RRID:AB_2864286) |
| PerCP/Cyanine5.5 Rat IgG1, κ Isotype Control (Clone: RTK2071) | Biolegend | ( Cat# 400425, RRID:AB_893689) |
| Anti-Mouse IgG (H+L) Cross-Adsorbed Secondary Antibody HRP coupled (Polyclonal) | Thermo Fisher Scientific | (Thermo Fisher Scientific Cat# G-21040, RRID:AB_2536527) |
| Anti-Mouse IgM Heavy Chain Antibody HRP Conjugated (Polyclonal) | Bethyl Laboratories | (Bethyl Cat# A90-101P, RRID:AB_67189) |
| Anti-mouse CD16/CD32 (Clone: 2.4G2) | BioXCell | (Bio X Cell Cat# BE0307, RRID:AB_2736987) |
| Anti-mouse CD3ε F(ab')2 fragment (Clone: 145-2C11) | BioXCell | (Bio X Cell Cat# BE0001-1FAB, RRID:AB_2687679) |
| Anti-mouse CD28 (Clone: PV-1) | BioXCell | (Bio X Cell Cat# BE0015-5, RRID:AB_1107628) |
| Anti-human CD3 (Clone: OKT3) | Thermo Fisher Scientific | (Thermo Fisher Scientific Cat# 16-0037-81, RRID:AB_468854) |

| | | |
|---|---|---|
| Anti-human/monkey CD28 (Clone: CD28.2) | BioXCell | (Bio X Cell Cat# BE0291, RRID:AB_2687814) |
| Purified Anti-human CD279 (PD-1) (Clone: EH12.2H7) | Biolegend | ( Cat# 329902, RRID:AB_940488) |
| Purified Mouse IgG1, κ Isotype Control (Clone: MOPC-21) | Biolegend | ( Cat# 400102, RRID:AB_2891079) |
| Purified Anti-human IL-6 (Clone: MQ2-39C3) | Biolegend | ( Cat# 501204, RRID:AB_2296206) |
| Purified Anti-human IL-4 Antibody (Clone: MP4-25D2) | Biolegend | ( Cat# 500802, RRID:AB_315121) |
| Purified Rat IgG1, κ Isotype Control (Clone: RTK2071) | Biolegend | ( Cat# 400402, RRID:AB_326508) |
| **Chemicals, biochemicals, biologics and proteins** | | |
| Diphtheria Toxin, Unnicked, Corynebacterium diphtheriae - Calbiochem | Sigma-Aldrich | Catalog # 322326 |
| DPBS, 10x, no calcium, no magnesium | Thermo Fisher Scientific | Catalog # 14200-067 |
| PBS, pH 7.2 | Thermo Fisher Scientific | Catalog # 20012-019 |
| Imject Alum Adjuvant | Thermo Fisher Scientific | Catalog # 77161 |
| Tween 20, 100% Nonionic Detergent | Bio-Rad | Catalog # 1706531; CAS: 9005-64-5 |
| Sm/RNP Complex Antigen affinity purified | ImmunoVision | Catalog # SRC-3000 |
| Deoxyribonucleic acid sodium salt from calf thymus | Sigma-Aldrich | Catalog # D1501; CAS: 73049-39-5 |
| Goat Serum Donor Herd | Sigma-Aldrich | Catalog # G6767 |
| Pierce DNA Coating Solution | Thermo Fisher Scientific | Catalog # 17250 |
| Albumin from chicken egg white | Sigma-Aldrich | Catalog # A5503; CAS: 9006-59-1 |
| Trypan Blue Solution, 0.4% | Thermo Fisher Scientific | Catalog # 15250-061 |
| RPMI 1640 Medium, GlutaMAX Supplement, HEPES | Thermo Fisher Scientific | Catalog # 72400-021 |
| MEM Non-Essential Amino Acids Solution (100X) | Thermo Fisher Scientific | Catalog # 11140-035 |
| Fetal Bovine Serum, qualified, Brazil | Thermo Fisher Scientific | Catalog # 10270-106; Lot: 2275142 |
| Penicillin - streptomycin (5 000 U/ml) | Thermo Fisher Scientific | Catalog # 15070-063 |
| Sodium Pyruvate (100 mM) | Thermo Fisher Scientific | Catalog # 11360-039 |
| 2-Mercaptoethanol | Sigma-Aldrich | Catalog # M6250-100ML; CAS: 60-24-2 |
| EDTA UltraPure 0.5M, pH de 8,0 | Thermo Fisher Scientific | Catalog # 15575-020 |
| Phorbol 12-myristate 13-acetate | Sigma-Aldrich | Catalog # P8139-1MG; CAS: 16561-29-8 |
| Ionomycin from Streptomyces conglobatus | Sigma-Aldrich | Catalog # I9657-1MG; CAS: 56092-81-0 |
| Brefeldin A | Sigma-Aldrich | Catalog # B7651; CAS: 20350-15-6 |
| Pristane | Sigma-Aldrich | Catalog # P2870; CAS: 1921-70-6 |
| Heparin sodium salt from porcine intestinal mucosa | Sigma-Aldrich | Catalog # H4784; CAS: 2608411 |
| Rat Sprague Dawley IgG Affinity Purified | Innovative Research | Catalog # IRTSDIGGAP50MG |
| Hamster Armenian IgG Affinity Purified | Innovative Research | Catalog # IHMARIGG50MG |
| Human IgG Affinity Purified | Innovative Research | Catalog # IHUIGGAP1000MG |
| Goat IgG Fractionated Purified Lyophilized | Innovative Research | Catalog # IGTIGGGFLY1GM |

| | | |
|---|---|---|
| OCT embedding cryoembedding Matrix | Thermo Fisher Scientific | Catalog # LAMB/OCT |
| Ghost Dye Violet 510 | Tonbo | Catalog # 13-0870-T100 |
| Streptavidin, Alexa Fluor 647 conjugate | Thermo Fisher Scientific | Catalog # S21374 |
| UltraComp eBeads Plus Compensation Beads | Thermo Fisher Scientific | Catalog # 01-3333-42 |
| MagniSort Streptavidin Negative Selection Beads | Thermo Fisher Scientific | Catalog # MSNB-6002-74 |
| Intracellular Staining Permeabilization Wash Buffer (10X) | Biolegend | Catalog # 421002 |
| Fixation Buffer | Biolegend | Catalog # 420801 |
| Recombinant Murine IL-3 | Peprotech | Catalog # 213-13 |
| Recombinant Human IL-3 (carrier-free) | Biolegend | Catalog # 578006 |
| **Commercial Kits** | | |
| eBioscience Foxp3 / Transcription Factor Staining Buffer Set | Thermo Fisher Scientific | Catalog # 00-5523-00 |
| EasySep Human Naïve CD4+ T Cell Isolation Kit | Stemcell Technologies | Catalog # 19555 |
| EasySep Human Basophil Isolation Kit | Stemcell Technologies | Catalog # 17969 |
| EasySep Mouse Naïve CD4+ T Cell Isolation Kit | Stemcell Technologies | Catalog # 19765 |
| **Experimental models: Organisms/strains** | | |
| Mouse: B6.129P2-Gt(ROSA)26$^{Sortm1(DTA)Lky/J}$ C57BL/6 | The Jackson Laboratory | RRID:IMSR_JAX:009669 |
| Mouse: Il4$^{flox/flox}$ C57BL/6 | Shibata et al. Proc Natl Acad Sci USA. 2018 Dec 18;115(51):13057-13062. | N/A |
| Mouse: Cd274loxP (PD-L1$^{flox/flox}$) C57BL/6 | Schwartz et al. J Exp Med. 2017 Sep 4;214(9):2507-2521. | N/A |
| Mouse: Il-6$^{flox/flox}$ C57BL/6 | Quintana et al. Brain Behav Immun. 2013 Jan;27(1):162-73. | N/A |
| Mouse: Mcpt8$^{Cre\text{-}tdTomato}$ C57BL/6 | Tchen et al. Front Immunol. 2022 Jun 29;13:900532. | N/A |
| Mouse: Mcpt8$^{DTR}$ C57BL/6 | Wada et al. J Clin Invest. 2010 Aug;120(8):2867-75. | N/A |
| Mouse: B6.129S4-Lyn$^{tm1Sor/J}$ C57BL/6 | The Jackson Laboratory | RRID:IMSR_JAX:003515 |
| Mouse: B6.129S4-Lyn$^{tm1Sor/J}$ Mcpt8$^{DTR}$ C57BL/6 | Pellefigues et al. Nat Commun. 2018 Feb 20;9(1):725. | N/A |

| **Softwares** | | |
|---|---|---|
| FlowJo v10.8.1 | Tree Star | FlowJo (RRID:SCR_00852) |
| BD FACSDiva Software v8.0 | BD Biosciences | BD FACSDiva Software (RRID:SCR_00145) |

| | | |
|---|---|---|
| ImageJ | NIH, Schneider et al. Nat Methods. 2012 Jul;9(7):671-5. | ImageJ (RRID:SCR_00307) |
| Graphpad Prism 9.4 | Graphpad | GraphPad Prism (RRID:SCR_00279) |
| CellSens Dimensions 1.4 (Build 8583) | Olympus | Olympus cellSens Software (RRID:SCR_01455) |
| **Other** | | |
| BD LSRFortessa X-20 Cell Analyzer | BD Biosciences | N/A |
| BD FACSMelody Cell Sorter | BD Biosciences | N/A |
| BD LSRFortessa Cell Analyzer | BD Biosciences | N/A |
| Leica DM IRB Inverted microscope | Leica | N/A |
| Video camera microscope attachment | Hamamatsu | ORCA-03G02 |
| Fluorescence microplate reader | TECAN | SPARK 10M |

Supplementary Figure S1

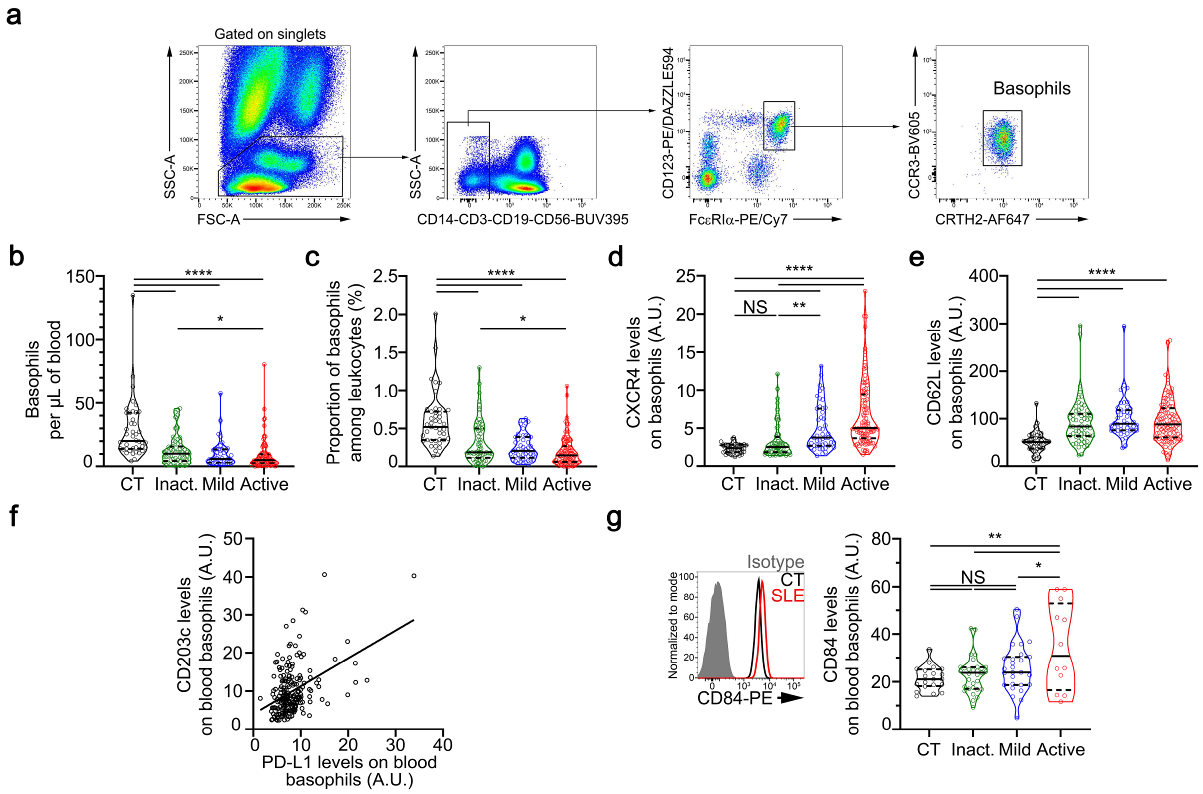

**Figure S1: Activated phenotype of basophils in SLE patients (related to Fig. 1)**

(**a**) Flow cytometry gating strategy used to define basophils in human blood samples. Basophils were defined as $SSC^{lo}$ $CD14^{-}$ $CD3^{-}$ $CD19^{-}$ $CD56^{-}$ $CD123^{+}$ $Fc\varepsilon RI\alpha^{+}$ $CCR3^{+}$ $CRTH2^{+}$ cells. (**b**) Absolute number of basophils per µL of blood in samples from healthy controls (CT) and inactive (inact.), mild or active SLE patients (n = 42/61/47/96, respectively) was determined by flow cytometry as described in (**a**). (**c**) Proportions (%) of basophils among leukocytes in blood samples as described in (**a**,**b**) (n = 43/61/47/98). (**d**) CXCR4 expression levels on blood basophils as described in (**a**,**b**) (n = 43/61/47/98, respectively) as determined by flow cytometry. (**e**) CD62L expression levels on blood basophils as described in (**a**,**b**) (n = 43/59/44/98, respectively) as determined by flow cytometry (**f**) Spearman correlation (and linear regression) between basophil CD203c and PD-L1 expression levels (r=0.3113, $P < 0.0001$, n=204). (**g**) **Left**, Representative FACS analysis of CD84 expression levels on blood basophils from a healthy control (CT, black line), a patient with active SLE (red line), and isotype control staining (grey filled histogram). **Right**, CD84 expression levels on blood basophils from healthy controls (CT) and inactive (inact.), mild or active SLE patients (n = 20/31/27/12, respectively) as determined by flow cytometry. (**b**-**e**, **g**) Data are presented as violin plots with median (plain line) and quartiles (dotted lines). Statistical analyses were Kruskal-Wallis tests followed by Dunn's multiple comparisons tests. NS: not significant, *$P < 0.05$, **$P < 0.01$, ****$P < 0.0001$. A.U. arbitrary units.

Supplementary Figure S2

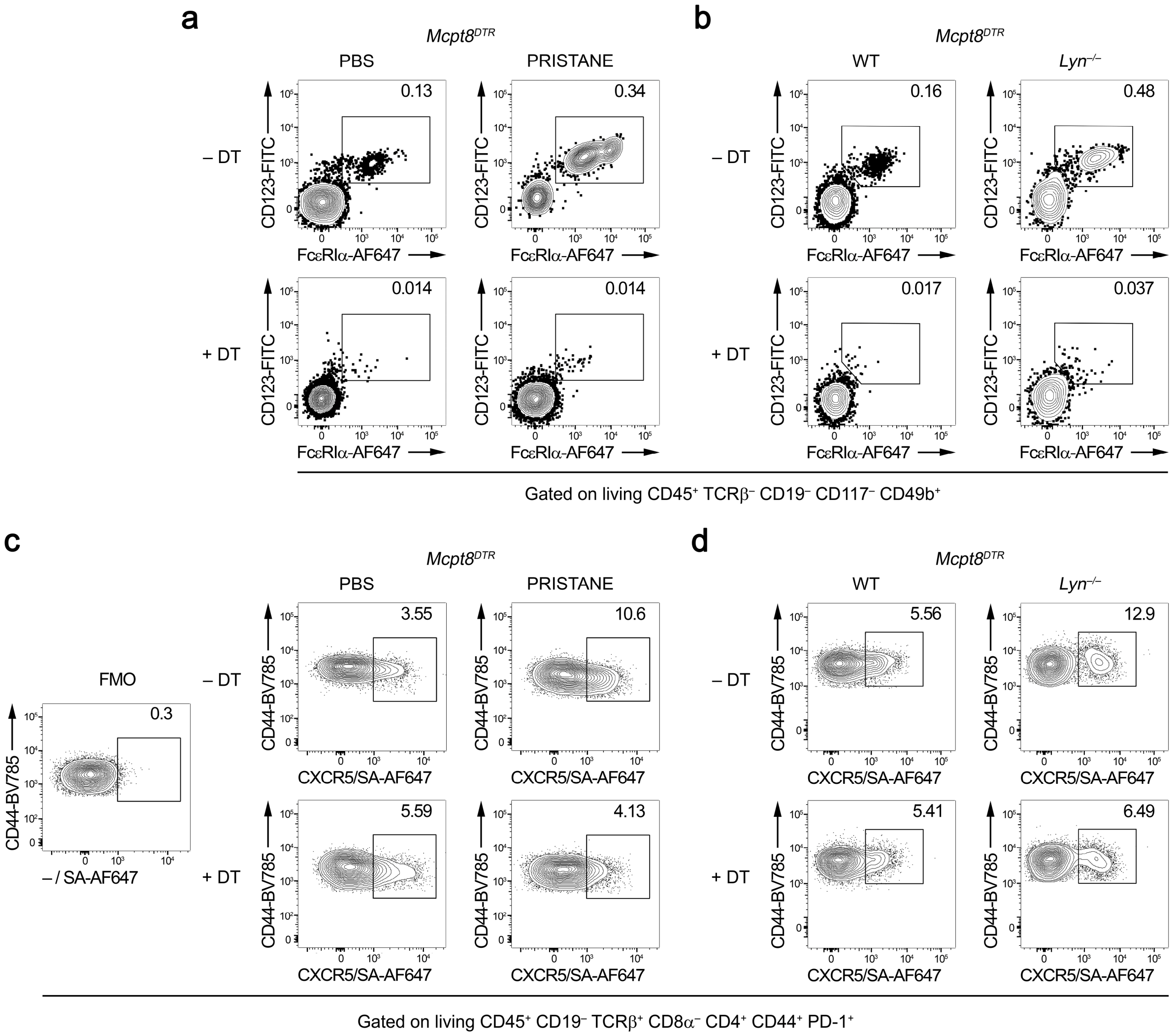

**Figure S2: Basophil and TFH gating strategy and effects of DT injection in lupus-like mouse models on *Mcpt8$^{DTR}$* background (related to Fig. 2)**

(**a**,**b**) Representative contour plots showing gating and proportions of basophils (defined as CD45$^+$ TCRβ$^-$ CD19$^-$ CD117$^-$ CD49b$^+$ CD123$^+$ FcεRIα$^+$ cells) among CD45$^+$ splenocytes in *Mcpt8$^{DTR}$* mice PBS- or pristane-injected (**a**) and in aged *Mcpt8$^{DTR}$* (WT) and *Lyn$^{-/-}$ Mcpt8$^{DTR}$* (*Lyn$^{-/-}$*) mice (**b**) basophil-depleted (+DT) or not (–DT). (**c**,**d**) Representative contour plots showing gating and proportions of TFH cells (defined as CD45$^+$ CD19$^-$ TCRβ$^+$ CD8α$^-$ CD4$^+$ CD44$^+$ PD-1$^+$ CXCR5$^+$ cells) among spleen CD4$^+$ T cells from mice as described in (**a**,**b**). CXCR5 positivity was determined on the fluorescence minus one (FMO, left panel). SA: streptavidin.

Supplementary Figure S3

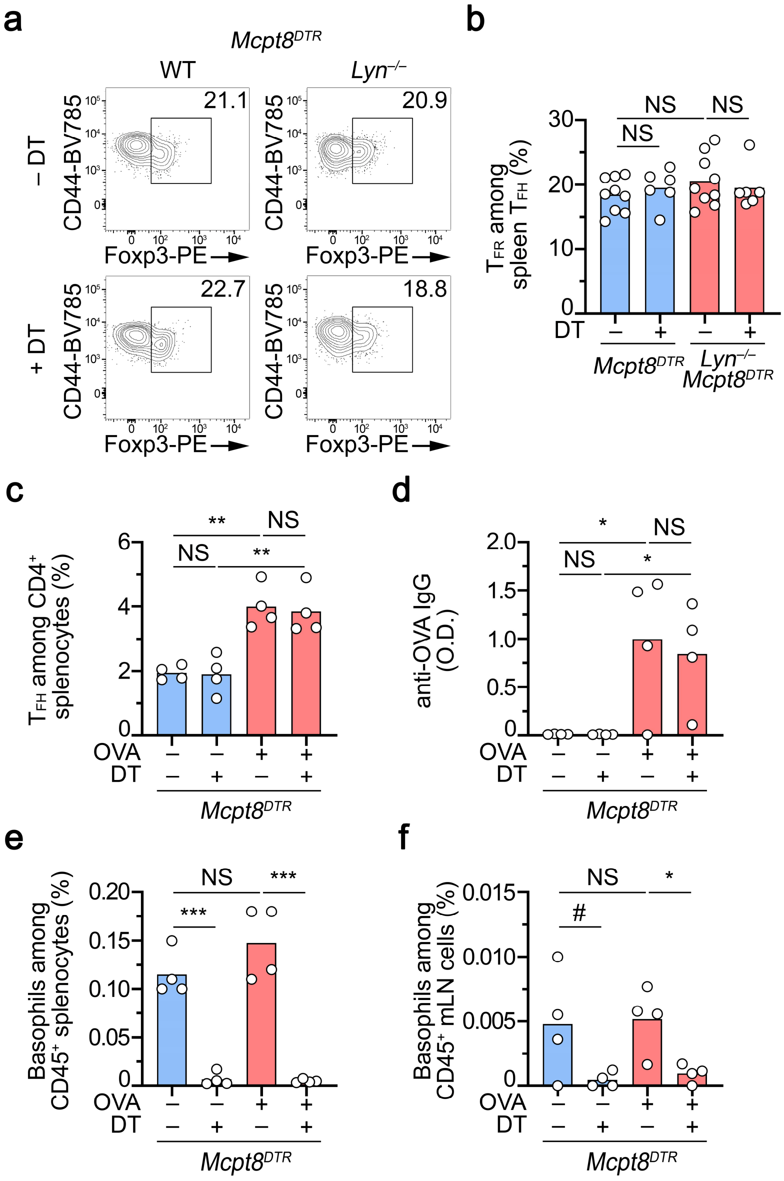

**Figure S3: Effects of basophil depletion on TFR/TFH ratio and ovalbumin immunization**

(**a**) Representative contour plots showing gating and proportions of regulatory TFH cells (TFR) (defined as Foxp3+ TFH cells) among spleen TFH cells (as described in **Fig. S2d**) from mice as described in **Fig. 2b**. (**b**) Proportions (%) of TFR among TFH cells as in (**a**). (**c**) Proportions (%) of TFH among $CD4^+$ T cells in $Mcpt8^{DTR}$ mice immunized with ovalbumin (OVA + ) or not (OVA –) and basophil-depleted (DT +) or not (DT –) 48 hours before analysis. (**d**) Anti-OVA IgG plasma levels in mice described in (**c**) were determined by ELISA. The values presented are 450 nm optical density values (O.D.). (**e**,**f**) Proportions (%) of basophils among $CD45^+$ splenocytes (**d**) and mesenteric lymph node (mLN) cells (**f**) of the mice described in (**c**). (**b**-**f**) Results are from at least three independent experiments and presented as individual values in bars representing the mean values. Statistical analyses were done by unpaired Student t-tests between the indicated groups. NS: not significant, $p>0.05$; #: $p=0.08$; *: $p<0.05$; **: $p<0.01$; ***: $p<0.001$.

Supplementary Figure S4

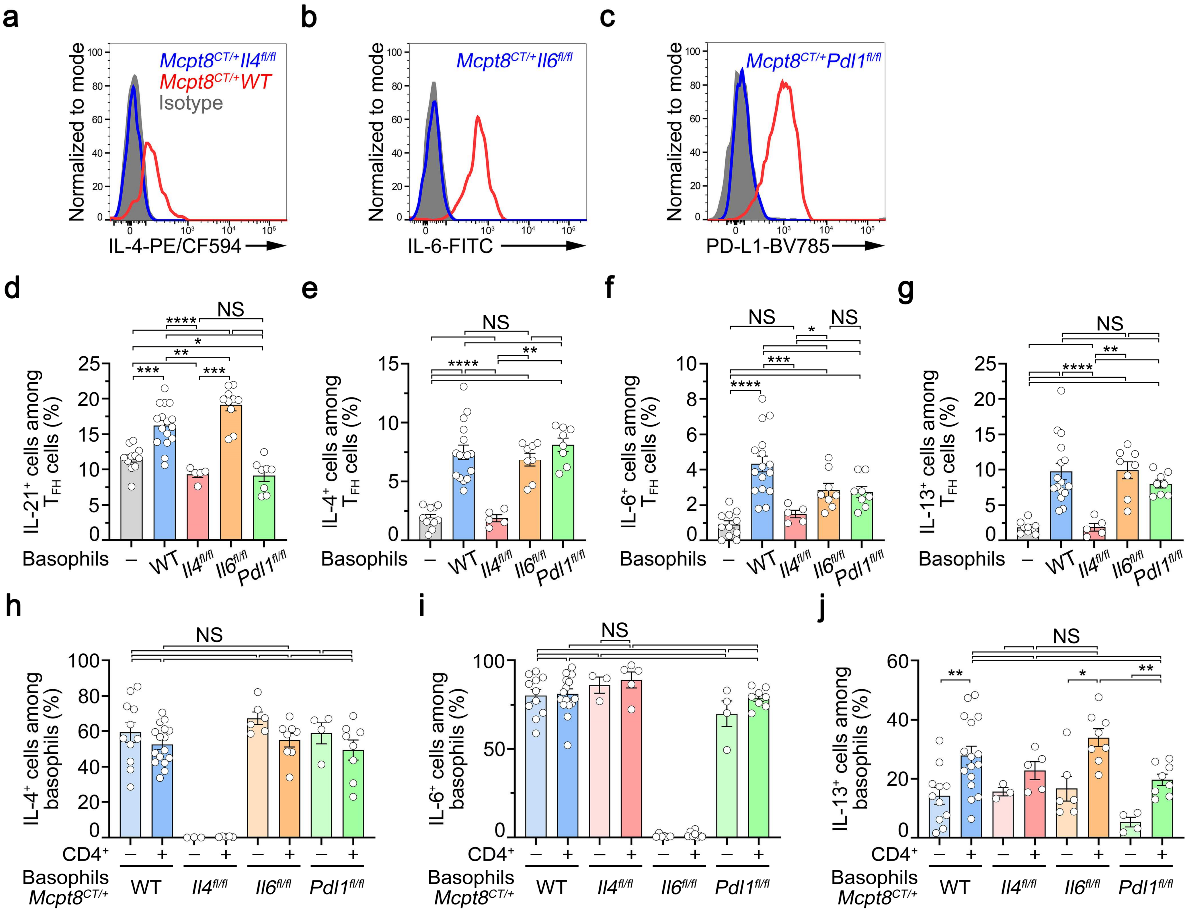

**Figure S4: Effective CRE-mediated floxed gene deletion in basophils and PMA-Ionomycin-induced cytokine production by basophils co-cultured or not with naïve CD4+ T cells (related to Fig. 4)**

(**a**) Representative FACS histogram plot showing IL-4 production by PMA and ionomycin stimulated spleen basophils from *Mcpt8*$^{CT/+}$ *Il4*$^{+/+}$ (WT, red line) and *Mcpt8*$^{CT/+}$ *Il4*$^{fl/fl}$ (IL-4 deficient basophils, blue line). The isotype control signal is overlaid (grey-filled histogram). (**b**) Representative FACS histogram plot showing IL-6 production by PMA and ionomycin stimulated spleen basophils from *Mcpt8*$^{CT/+}$ *Il6*$^{+/+}$ (WT, red line) and *Mcpt8*$^{CT/+}$ *Il6*$^{fl/fl}$ (IL-6 deficient basophils, blue line). The isotype control signal is overlaid (grey-filled histogram). (**c**) Representative FACS histogram plot showing PD-L1 expression on spleen basophils from *Mcpt8*$^{CT/+}$ *Pdl1*$^{+/+}$ (WT, red line) and *Mcpt8*$^{CT/+}$ *Pdl1*$^{fl/fl}$ (PD-L1 deficient basophils, blue line). The isotype control signal is overlaid (grey-filled histogram). (**d**-**g**) Proportions (%) of IL-21- (**d**), IL-4- (**e**), IL-6- (**f**), and IL-13- (**g**) producing cells among TFH cells restimulated with PMA and ionomycin and brefeldin A for the last 4 hours of the culture from the same conditions as described in **Fig. 4**. (**h**-**j**) Proportions (%) of PMA and ionomycin-induced IL-4- (**d**), IL-6- (**e**) and IL-13- (**f**) producing cells among basophils of the indicated genotypes cultured without (–, light colors) or with (+, dark colors) purified wild-type CD3/CD28 activated naïve CD4+ T cells as described in **Fig. 4**. (**d**-**j**) Results are from at least three independent experiments and presented as individual values in bars representing the mean values ± s.e.m. Statistical analyses were done by Mann-Whitney U tests between the indicated groups. NS: not significant, p>0.05; *: p<0.05; **: p<0.01.

Supplementary Figure S5

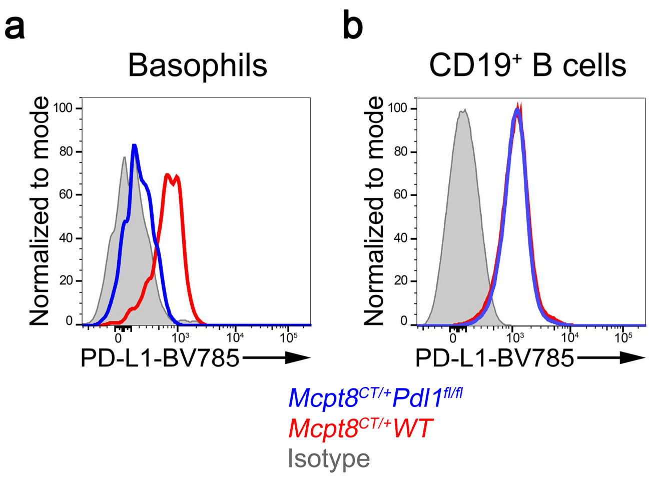

**Figure S5: CRE-mediated *Pdl1* floxed gene deletion selectively in the basophil compartment**

(**a**) Representative FACS histogram plot showing PD-L1 expression on spleen basophils from *Mcpt8*$^{CT/+}$ *Pdl1*$^{+/+}$ (WT, red line) and *Mcpt8*$^{CT/+}$ *Pdl1*$^{fl/fl}$ (PD-L1 deficient basophils, blue line). The isotype control signal is overlaid (grey-filled histogram). (**b**) Representative FACS histogram plot showing PD-L1 expression on spleen CD19$^{+}$ cells from the mice as in (**a**).

Supplementary Figure S6

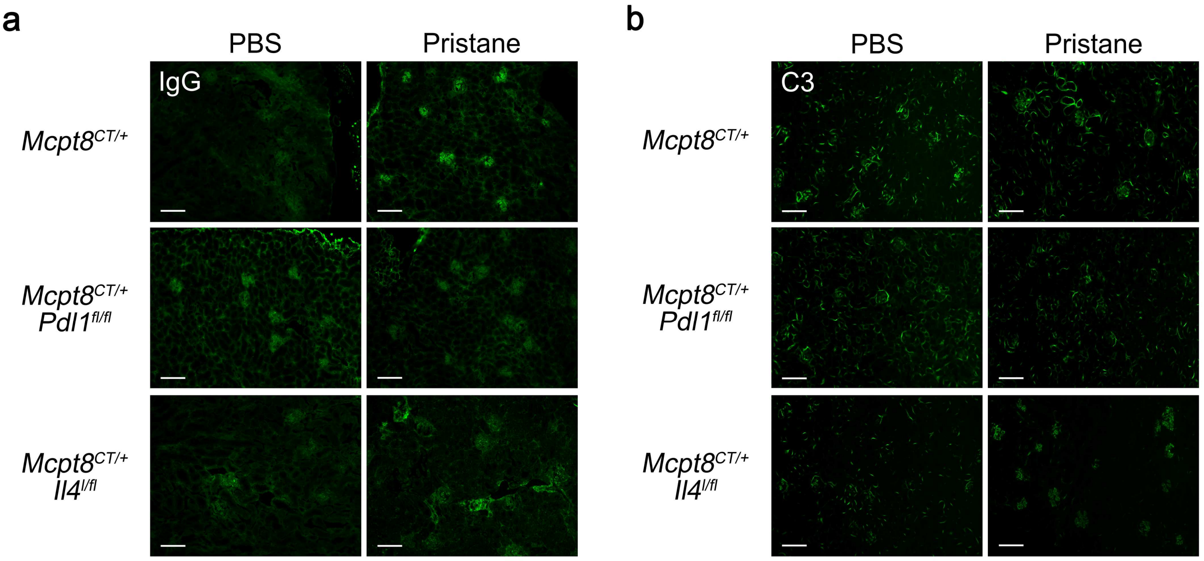

**Figure S6: IgG and C3 deposits in kidneys of PBS- or pristane-injected animals (related to Figures 5 and 6)**

Representative pictures of one field of kidney from mice with the indicated genotypes treated without (PBS) or with pristane for 8 weeks showing the intensity of anti-IgG (**a**) or anti-C3 (**b**) staining by immunofluorescence. Scale bar = 200 µm.
